# Diagnosis of COVID-19 Using CT image Radiomics Features: A Comprehensive Machine Learning Study Involving 26,307 Patients

**DOI:** 10.1101/2021.12.07.21267367

**Authors:** Isaac Shiri, Yazdan Salimi, Abdollah Saberi, Masoumeh Pakbin, Ghasem Hajianfar, Atlas Haddadi Avval, Amirhossein Sanaat, Azadeh Akhavanallaf, Shayan Mostafaei, Zahra Mansouri, Dariush Askari, Mohammadreza Ghasemian, Ehsan Sharifipour, Saleh Sandoughdaran, Ahmad Sohrabi, Elham Sadati, Somayeh Livani, Pooya Iranpour, Shahriar Kolahi, Bardia Khosravi, Maziar Khateri, Salar Bijari, Mohammad Reza Atashzar, Sajad P. Shayesteh, Mohammad Reza Babaei, Elnaz Jenabi, Mohammad Hasanian, Alireza Shahhamzeh, Seyed Yaser Foroghi Gholami, Abolfazl Mozafari, Hesamaddin Shirzad-Aski, Fatemeh Movaseghi, Rama Bozorgmehr, Neda Goharpey, Hamid Abdollahi, Parham Geramifar, Amir Reza Radmard, Hossein Arabi, Kiara Rezaei-Kalantari, Mehrdad Oveisi, Arman Rahmim, Habib Zaidi

**Author notes:** Corresponding author: Habib Zaidi, Ph.D, Geneva University Hospital, Division of Nuclear Medicine and Molecular Imaging CH-1211 Geneva, Switzerland. First Author: Isaac Shiri, MSc, Geneva University Hospital, Division of Nuclear Medicine and Molecular Imaging, CH-1211 Geneva, Switzerland.

## Abstract

**Purpose:** To derive and validate an effective radiomics-based model for differentiation of COVID-19 pneumonia from other lung diseases using a very large cohort of patients.

**Methods:** We collected 19 private and 5 public datasets, accumulating to 26,307 individual patient images (15,148 COVID-19; 9,657 with other lung diseases e.g. non-COVID-19 pneumonia, lung cancer, pulmonary embolism; 1502 normal cases). Images were automatically segmented using a validated deep learning (DL) model and the results carefully reviewed. Images were first cropped into lung-only region boxes, then resized to 296×216 voxels. Voxel dimensions was resized to 1×1×1mm^3^ followed by 64-bin discretization. The 108 extracted features included shape, first-order histogram and texture features. Univariate analysis was first performed using simple logistic regression. The thresholds were fixed in the training set and then evaluation performed on the test set. False discovery rate (FDR) correction was applied to the p-values. Z-Score normalization was applied to all features. For multivariate analysis, features with high correlation (R^2^>0.99) were eliminated first using Pearson correlation. We tested 96 different machine learning strategies through cross-combining 4 feature selectors or 8 dimensionality reduction techniques with 8 classifiers. We trained and evaluated our models using 3 different datasets: 1) the entire dataset (26,307 patients: 15,148 COVID-19; 11,159 non-COVID-19); 2) excluding normal patients in non-COVID-19, and including only RT-PCR positive COVID-19 cases in the COVID-19 class (20,697 patients including 12,419 COVID-19, and 8,278 non-COVID-19)); 3) including only non-COVID-19 pneumonia patients and a random sample of COVID-19 patients (5,582 patients: 3,000 COVID-19, and 2,582 non-COVID-19) to provide balanced classes. Subsequently, each of these 3 datasets were randomly split into 70% and 30% for training and testing, respectively. All various steps, including feature preprocessing, feature selection, and classification, were performed separately in each dataset. Classification algorithms were optimized during training using grid search algorithms. The best models were chosen by a one-standard-deviation rule in 10-fold cross-validation and then were evaluated on the test sets.

**Results:** In dataset #1, Relief feature selection and RF classifier combination resulted in the highest performance (Area under the receiver operating characteristic curve (AUC) = 0.99, sensitivity = 0.98, specificity = 0.94, accuracy = 0.96, positive predictive value (PPV) = 0.96, and negative predicted value (NPV) = 0.96). In dataset #2, Recursive Feature Elimination (RFE) feature selection and Random Forest (RF) classifier combination resulted in the highest performance (AUC = 0.99, sensitivity = 0.98, specificity = 0.95, accuracy = 0.97, PPV = 0.96, and NPV = 0.98). In dataset #3, the ANOVA feature selection and RF classifier combination resulted in the highest performance (AUC = 0.98, sensitivity = 0.96, specificity = 0.93, accuracy = 0.94, PPV = 0.93, NPV = 0.96).

**Conclusion:** Radiomic features extracted from entire lung combined with machine learning algorithms can enable very effective, routine diagnosis of COVID-19 pneumonia from CT images without the use of any other diagnostic test.

## INTRODUCTION

The recent pandemic caused by severe acute respiratory syndrome coronavirus 2 (SARS-CoV-2) has raised great concerns worldwide ^1^. As of December 2021, it has been responsible for more than 265 million confirmed cases and 5 million deaths all over the globe (see who.int). Different diagnostic methods have been proposed for SARS-CoV-2, also known as COVID-19. The most popular is the real-time reverse transcriptase-polymerase chain reaction (RT-PCR), which is a molecular test ^2–6^. Although RT-PCR is a highly reputable diagnostic test, it has some drawbacks, such as the delay in providing test reports, the remarkable effort of healthcare professionals in the process of sampling, and last but not least, the limited availability of the test kits especially in developing countries. In this regard, many physicians have turned to other diagnostic methods, including chest X-ray (CXR) and computed tomography (CT). CXR plays an important role in the non-invasive, fast, widely available, and cost-effective assessment of pulmonary lesions ^7^. In the context of COVID-19, some characteristics, such as ground glass or patchy opacities can be linked with COVID-19 pneumonia in a CXR. Nonetheless, discovering these findings requires expertise as these changes are subtle ^8^. CT images, on the other hand, better demonstrate the opacities and are useful even for the early diagnosis of COVID-19 pneumonia in asymptomatic/pre-symptomatic patients ^9^. Moreover, CT shows a significantly higher sensitivity compared to CXR ^10, 11^. Thus, it is better suited in the triage of patients if a scanner is available and the higher dose of X-rays to the patient via CT imaging is carefully considered ^11^.

A wide range of studies has been published on the clinical use of CT and its qualitative and quantitative analysis for the diagnosis and management of COVID-19 patients ^12, 13^. A number of qualitative/semi-quantitative findings in chest CT, such as the presence and laterality of ground-glass opacities and consolidation, number of lobes affected, the extent of lung involvement are used as acceptable features for COVID-19 diagnosis ^14^. Nevertheless, these findings are most likely to be subjective, rely on physician’s inference, and have very low specificity for a definitive diagnosis of COVID-19 ^15^. Artificial Intelligence (AI) has been proposed as a solution to address the aforementioned limitations of CT (e.g., low specificity) as it provides with tools that can visualize and extract the most subtle and minute characteristics of images.

Radiomics, a high-level image analysis technique that mines multi-dimensional data from medical images, has especially emerged in the past decade ^16–30^ and is continuing to grow. It has been utilized in a range of diseases towards improved diagnosis, prognosis, treatment response assessment and other aspects of patient management. Recently, COVID-19 researchers have conducted a number of CT radiomics studies ^17–19, 31^. For prognosis, Tang et al. ^32^ focused on the prediction of COVID-19 severity using radiomics features of CT images and laboratory data. In another study, Wu et al. ^33^ analyzed the prognostic ability of radiomics features based on CT images of 492 patients to determine the poor-outcome group. Studies reported on the feasibility of deep learning (DL)/radiomics applied to CT images towards classification (e.g. COVID-19 pneumonia, non-COVID-19 pneumonia, other lung diseases, or normal images). Harmon et al. ^34^ trained a DL-based neural network and validated it on a cohort of 1337 patients. Bai et al. ^35^ included 1186 patients and evaluated an AI model for classifying CT images into COVID-19 and non-COVID-19 pneumonia. Zhang et al. ^36^ conducted a study on 3777 patients and evaluated an AI system with the addition of clinical data to determine whether an image reflects COVID-19 pneumonia, other pneumonia, or normal. Di et al. ^37^ utilized radiomic features to assess the differentiation of COVID-19 pneumonia and community-acquired pneumonia in 3330 patients. Xie et al. ^38^ evaluated the ability of radiomics features and ground-glass opacities in 301 patients for the discrimination of COVID-19 and non-COVID-19 patients.

Most aforementioned studies utilized datasets consisting of the dichotomy of COVID-19 pneumonia vs. other non-COVID-19 pneumonia, or COVID-19 vs. normal patients. However, some CXR or CT studies included other lung diseases as well. For instance, Albahli et al. ^39^ included 14 classes of diseases (e.g. cancer, pneumothorax, fibrosis, edema, atelectasis, etc.) in their dataset and trained their model on CXR images. Das et al. ^40^ also included tuberculosis patients and added their images to COVID-19 pneumonia, other pneumonia, and healthy CXR images of their dataset. Wang et al. ^41^ studied a dataset consisting of COVID-19 and other lung disease images in addition to normal lung images.

Several limitations were reported in a systematic review by Roberts et al. ^42^, which included more than 2,000 original articles focusing on the development of different DL/ML-based algorithms in diagnosis/prognosis of COVID-19 patients. First and foremost is data bias, many articles have used datasets with small sample sizes, duplicate samples, low quality or non-standardized medical image format. Moreover, many researchers have studied so-called Frankenstein ^42^ and Toy ^43^ datasets, utilizing small and/or low-quality images, which assemble and redistributed from other datasets ^42^. In addition, most of studies have not provided sufficient information regarding data preprocessing, demographics of training/testing cohorts, and code and data availability. Roberts et al. ^42^ reported that most articles in the diagnostic era failed to balance the number of COVID-19 and other classes of diseases in the training and testing datasets. For example, a study may have included considerably fewer COVID-19 cases compared to other cases. Regarding the methodology, most of the studies failed to elucidate an exact methodology which is a must for conducting a reproducible study. Hence, few studies are practical in real clinical situations.

In the present study, we have gathered a multi-institutional, multi-national CT image datasets of more than 26,307 patients containing COVID-19, non-COVID-19 pneumonia, and other lung pathologies. We aimed to assess the value of systematically utilizing CT-based radiomics to distinguish COVID-19 pneumonia from non-COVID-19 pneumonia and other types of lung diseases, using multiple dimensionality reduction, feature selection, and classification algorithms. To this end, we have assembled and utilized a very large, curated dataset and applied different AI, radiomics and prognostic/diagnostic modeling guidelines.

## MATERIALS AND METHODS

The methodological steps adopted in this study can be found in Figure 1. We filled out several standardized checklists regarding diagnostic modelling (TRIPOD, Transparent Reporting of a multivariable prediction model for Individual Prognosis Or Diagnosis [40]), and AI in medical image analysis (CLAIM, Checklist for Artificial Intelligence in Medical imaging ^44^) to ensure the reproducibility and decency of our study. The two checklists were completed separately by two expert medical imaging scientists in the field of radiomics and AI (with consensus) who were not co-authors of this study. The complete checklist of standardizations was provided in the supplementary files.

**Figure 1.**
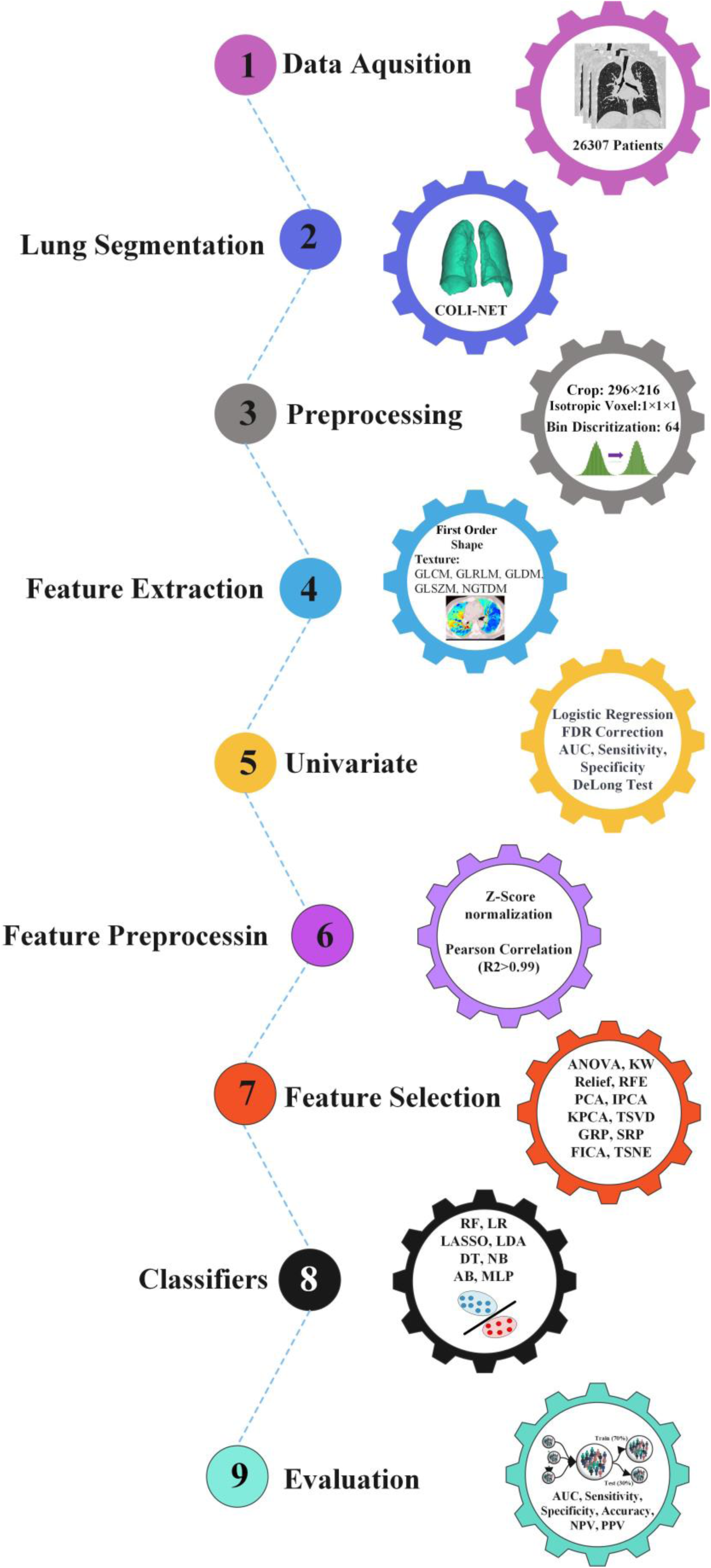
Flowchart of different steps implemented in this study.

### Datasets

The data of this study consisted of 18 local and 5 public datasets containing both COVID-19 and other lung diseases, arriving at 26,307 individual patient images (15,148 COVID-19; 9,657 with other lung diseases including non-COVID-19 pneumonia, lung cancer pulmonary embolism; and 1,502 normal). Our public dataset consisted of 5 separate datasets, including 1,744 COVID-19 ^34, 45, 46^, pulmonary embolism (PE, n=5,696) ^47^, and lung cancer (n=1,379) ^45^ CT images. Our private datasets were assembled in this effort from 18 clinical centers in Iran totaling 13,404 COVID-19 patients with the same number of CT images (one image per patient). Our study was approved by the local ethics committees, and written informed consent was waived owing to its retrospective nature.

To be included in the study, patients had to have either a positive RT-PCR result or positive CT findings. CT findings were considered as consistent with COVID-19 if: (a) typical COVID-19 patterns and manifestations are observed as described in the COVID-19 Reporting and Data System (CO-RADS) ^48^, and (b) two radiologists separately reached a conclusion that CT images are compatible with COVID-19 patterns, and (c) a third radiologist confirmed the diagnosis if the two former radiologists face a discrepancy in their opinions. After including the eligible patients, we applied our exclusion criteria to prepare a cleaner dataset. The exclusion criteria were as follows: patients who had negative RT-PCR result (n= 1,900), patients with severe motion artefacts in CT images confirmed by an experienced medical physicist (n= 560), patients with inappropriate positions in CT images (n= 121), patients with low-quality CT images (n= 210). In addition, 1,379 normal cases without any sign in CT images or background of lung disease, and 2,582 patients with other pneumonia (non-COVID-19) were enrolled to this study. After applying the aforementioned exclusion criteria, a total of 26,307 patients were enrolled as our overall dataset. Images were acquired in different medical centers using various CT scanners, there was a variability in acquisition parameters such as tube current and slice thickness. In our local medical centers, CT imaging was performed at the end inhalation breath-hold to decrease respiratory motion artefacts. More information on public database were provided in ^34, 45–47^.

### Image Segmentation, Image Preprocessing, and Feature Extraction

The images were segmented automatically using a previously constructed DL algorithm ^49^ and were reviewed to confirm the segmentation outcome. The images were first cropped to attain the lung-only region box, then resized to 296×216. Image voxel size was resized to 1×1×1 mm^3^ followed by 64-bin discretization. Radiomics feature extraction was performed using the Pyradiomcis library ^50^, which has been standardized according to the Image Biomarker Standardization Initiative (IBSI) ^51^. The 108 extracted features included shape (n=16), first-order histogram, second-order Gray Level Co-occurrence Matrix (GLCM, n=24), and higher-order features including Gray Level Dependence Matrix (GLDM, n=14), Gray Level Size Zone Matrix (GLSZM, n=16), Gray Level Run Length Matrix (GLRLM, n=16), and Neighboring Gray Tone Difference Matrix (NGTDM, n=5).

### Univariate Analysis

Univariate analysis was performed by using simple logistic regression; each feature was trained on the training sets, and the results were reported on the testing sets. Benjamin and Hochberg false discovery rate (FDR) corrections was applied to the p-values. Statistical comparison of AUCs between training sets and test sets was performed by the DeLong test [41] to ascertain best performance.

### Feature Preprocessing, Feature Selection, and Classifiers

Z-Score normalization was applied to all features. The mean and standard deviation were calculated in the training sets and then applied to testing sets. Features with high correlation (R^2^>0.99) were eliminated using Pearson correlation. In this study, we used 4 feature selection algorithms, including Analysis of Variance (ANOVA), Kruskal-Wallis (KW), Recursive Feature Elimination (RFE), and Relief, and 8 dimensionality reduction techniques, including Principal Component Analysis (PCA), Incremental PCA (IPCA), Kernel PCA (KPCA), Truncated SVD (TSVD), Gaussian Random Projection (GRP), Sparse Random Projection (SRP), Fast ICA (FICA), and TSNE. For the classification task, we used 8 classifiers, including Logistic Regression (LR), Least Absolute Shrinkage and Selection Operator (LASSO), Linear Discriminant Analysis (LDA), Decision Tree (DT), Random Forest (RF), AdaBoost (AB), Naïve Bayes (NB), and Multilayer Perceptron (MLP). By cross-combining our 4 feature selectors + 8 dimensionality reduction techniques with the 8 classifiers, we tested 96 different combinations.

### Evaluation

We trained and evaluated our models in 3 different scenarios. First of all, the entire dataset (26,307 patients including 15,148 COVID-19 and 11,159 non-COVID-19) being randomly split into 70% (18,414 patients) and 30% (7,893 patients) for the training and test sets, respectively. As our data included patients whose COVID-19 was confirmed using RT-PCR and patients confirmed only by imaging, this dataset included both populations. Second, excluding normal patients in other lung disease class, and only including RT-PCR positive COVID-19 cases in COVID-19 class, the resulting dataset (20,697 patients including 12,419 COVID-19, and 8,278 non-COVID-19) was randomly split into 70% (14,677 patients) and 30% (6,209 patients) for the training and test sets, respectively. In Third dataset, only including non-COVID-19 pneumonia patients and a random sample of COVID-19 patients (5,582 patients including 3,000 COVID-19 and 2,582 non-COVID-19) to provide a balanced dataset, and then randomly split the dataset into 70% (3,907 patients) and 30% (1,675 patients) for the training and test sets, respectively.

The multivariate steps, including feature preprocessing, feature selection, and classification were performed separately for each dataset. Classification algorithms were optimized during training using grid search algorithms. The best models were chosen by a one-standard-deviation rule in 10-fold cross-validation and then evaluated on test sets. The accuracy, sensitivity, specificity, area under the receiver operating characteristic curve (AUC), positive predictive value (PPV), and negative predicted value (NPV) were reported for the test set. Statistical comparison of AUCs between the different models in the test sets was performed by the DeLong test ^52^ to ascertain best performances. The significance level was considered at the level of 0.05. All multivariate analysis steps were performed in the Python open-source library Scikit-Learn ^53^.

## RESULTS

Figure 2 shows the hierarchical clustering heat map of the radiomics feature distribution for COVID-19 and lung disease classes for the entire dataset (i.e. dataset #1). We provide a hierarchical clustering heat map for strategies 2 and 3 in Supplemental Figures 1 and 2, respectively. The correlation of radiomic features are depicted in Figure 3 for the entire dataset, whereas Supplemental Figures 3 and 4 show the same for strategies 2 and 3. Highly correlated features (R^2^>0.99) were identified and redundant features eliminated, enabling dimensionality reduction in an unsupervised manner, prior to multivariate analysis using machine learning algorithms.

**Figure 2.**
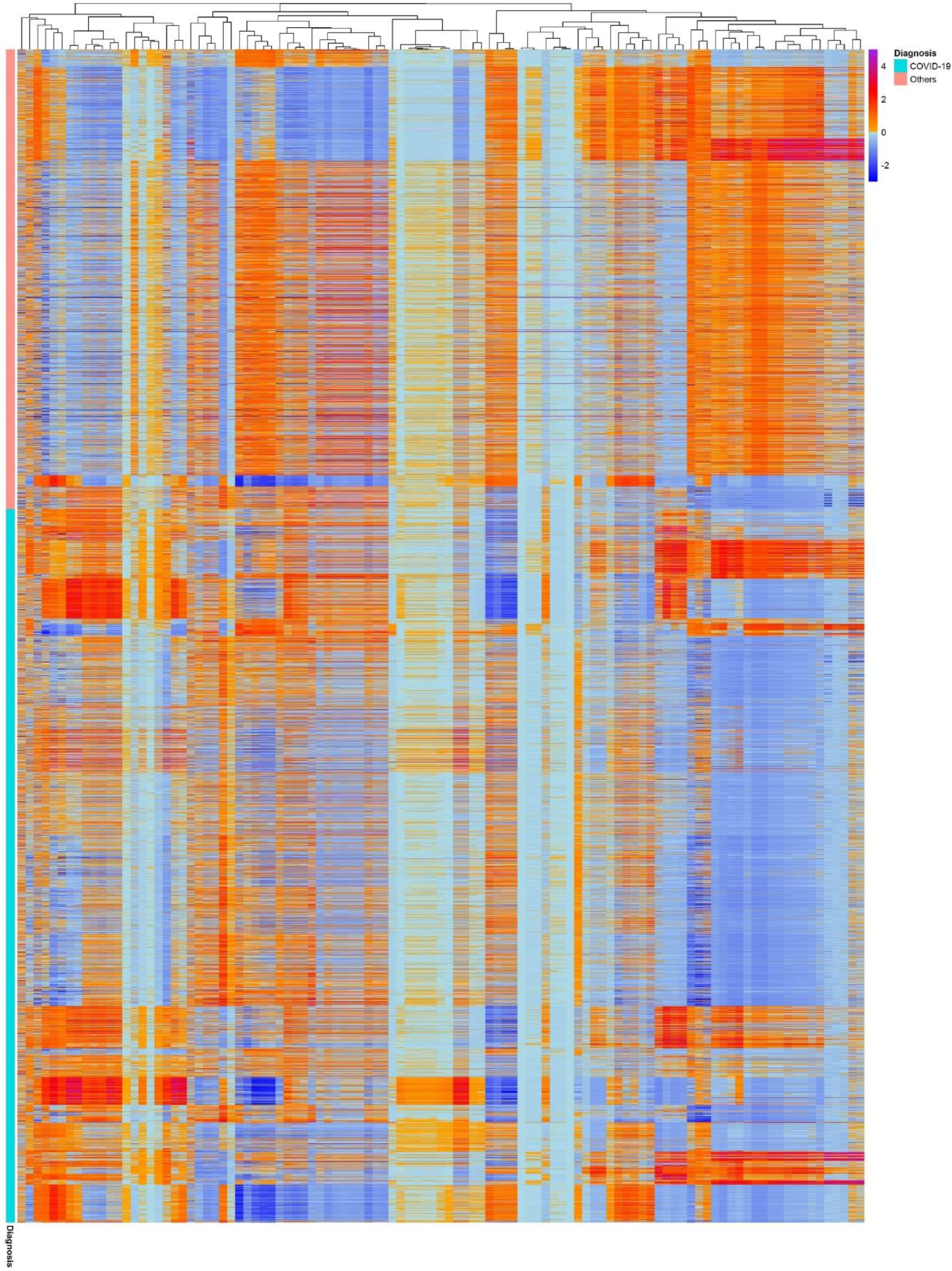
Cluster heat map of the whole dataset. Columns and rows are the radiomics features and patients sample, respectively.

**Figure 3.**
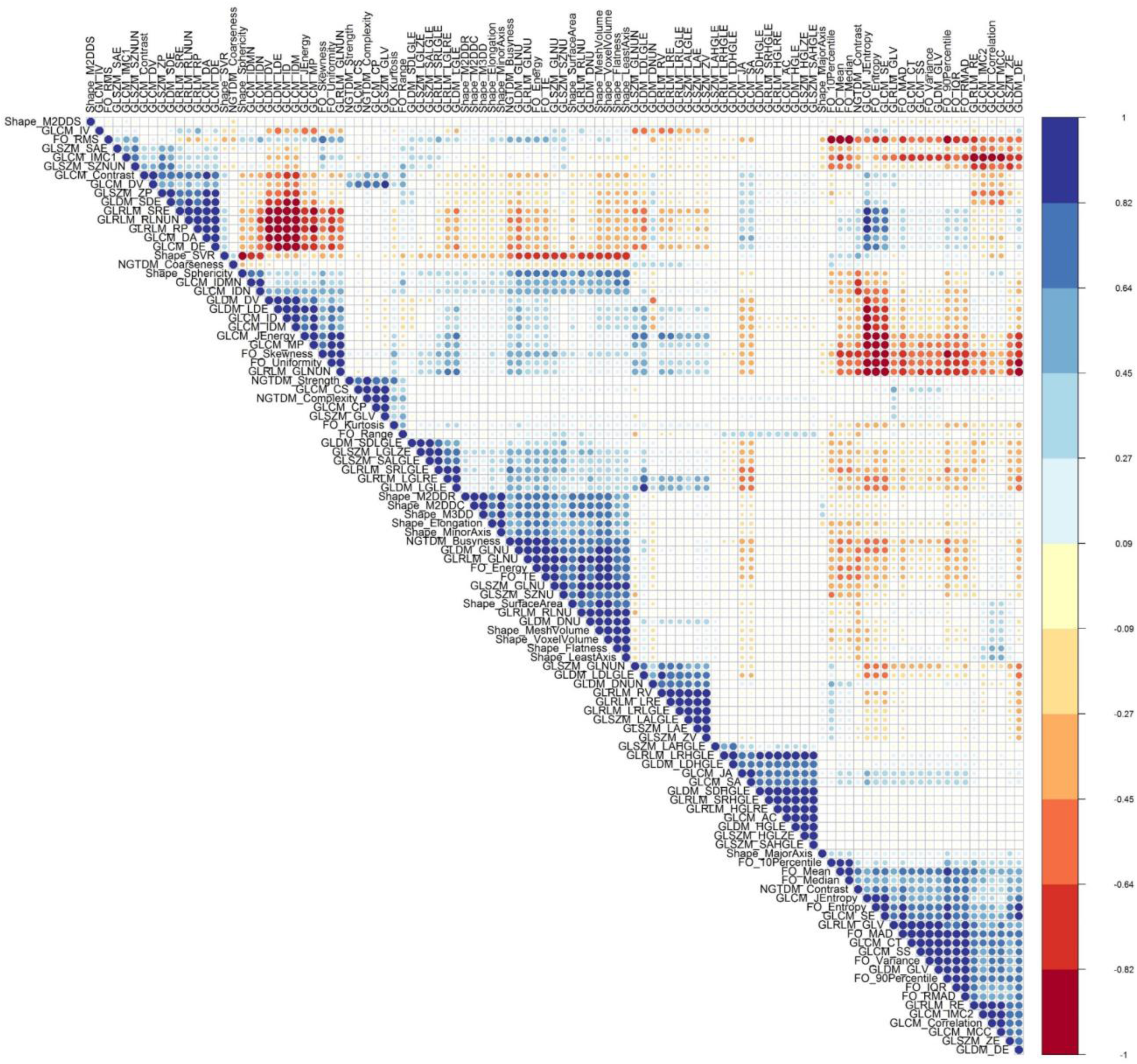
Correlation matrix between different features in the entire dataset.

Supplemental Tables 1-3 summarize univariate analysis for each feature in strategies 1, 2 and 3, respectively. We report the training and test AUC, sensitivity, specificity, and also adjusted p-values (using Benjamin and Hochberg FDR method) for each feature using binary logistic regression. When comparing AUCs, none of the features had a p-value<0.05 for the training and test sets using the DeLong test. In dataset #1, 3 features including Robust Mean Absolute Deviation from first-order (AUC = 0.55, Sensitivity = 0.54, Specificity = 0.56), Small Area High Gray Level Emphasis from GLSZM (AUC = 0.55, Sensitivity = 0.61, Specificity = 0.55), and Long-Run High Gray Level Emphasis form GLRLM (AUC = 0.5, Sensitivity = 0.54, Specificity = 0.50) had AUC, sensitivity and specificity higher than 0.50. For strategies 2 and 3, Robust Mean Absolute Deviation from first-order (AUC = 0.55, Sensitivity = 0.54 and Specificity = 0.51) and Large Dependence Emphasis from GLDM (AUC = 0.55, Sensitivity = 0.50 and Specificity = 0.57) had AUC, sensitivity and specificity higher than 0.5, respectively.

For dataset #1, Figure 4 represents the heatmap of the cross-combination of feature selectors and classifiers for AUC, sensitivity, specificity, accuracy, PPV, NPV, wherein collective datasets was randomly split into 70% (18,208 patients) and 30% (7,804 patients) for the training and test sets, respectively. Relief feature selection and RF classifier combination resulted in highest performance (AUC = 0.99, sensitivity = 0.98, specificity = 0.94, accuracy = 0.96, PPV = 0.96, NPV = 0.96). Supplemental Figure 5 presents a box plot of feature selectors and classifiers for the different evaluation metrics in dataset 1.

**Figure 4.**
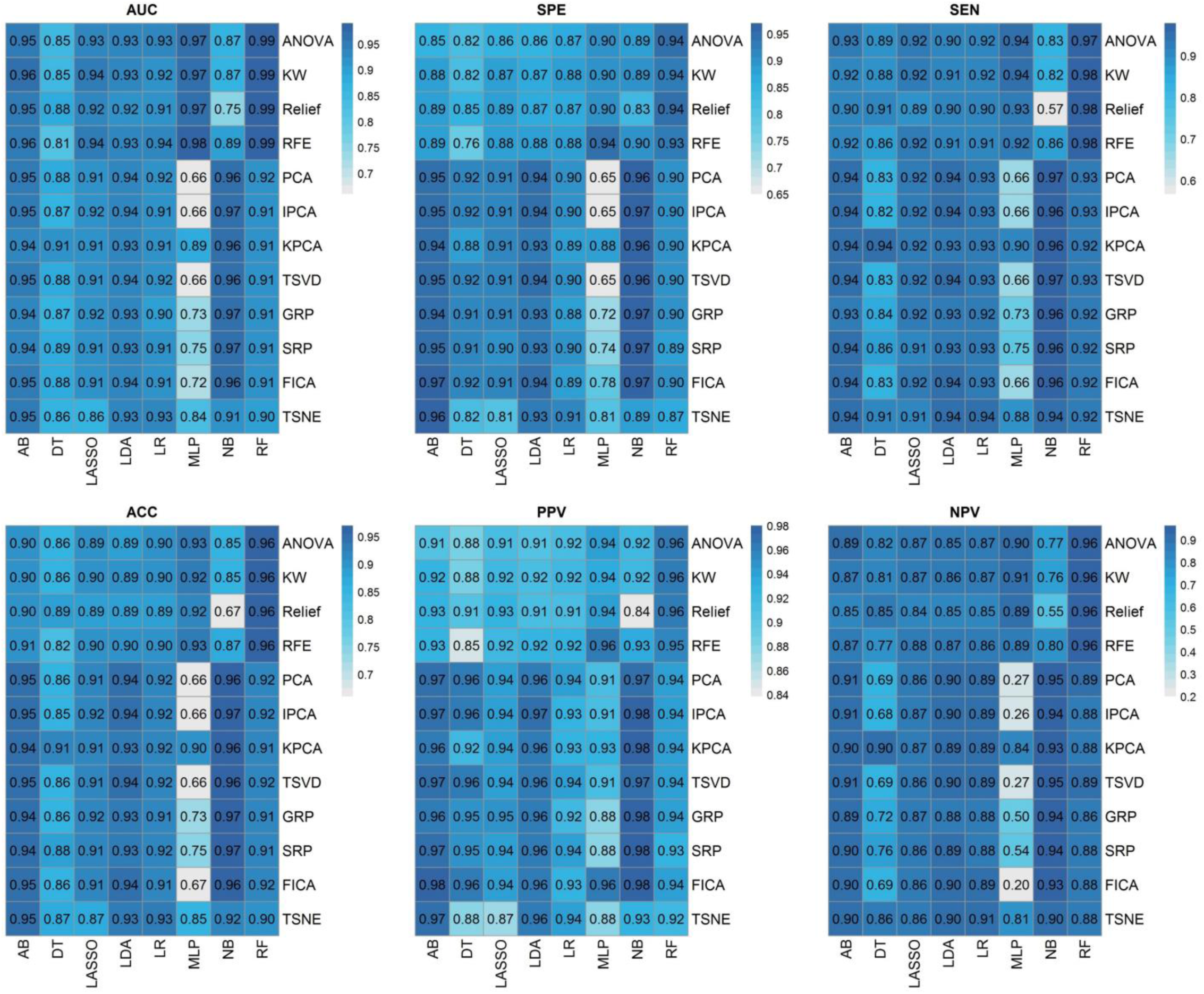
Heatmaps of the cross-combination of feature selectors (12 rows) and classifiers (8 columns) for AUC, sensitivity, specificity, accuracy, PPV, NPV in dataset #1.

For dataset #2, Figure 5 represents the heatmap of the cross-combination of feature selectors and classifiers for AUC, sensitivity, specificity, accuracy, PPV, NPV, wherein normal cases were excluded and CT images with PCR-Positive were included and then randomly split into 70% (15,514 patients) and 30% (6,649 patients) for the training and test sets, respectively. RFE feature selection and RF classifier combination resulted in highest performance (AUC = 0.99, sensitivity = 0.98, specificity = 0.95, accuracy = 0.97, PPV = 0.96, NPV = 0.98). Supplemental Figure 6 represents a box plot of feature selectors and classifiers for different evaluation metrics in dataset #2.

**Figure 5.**
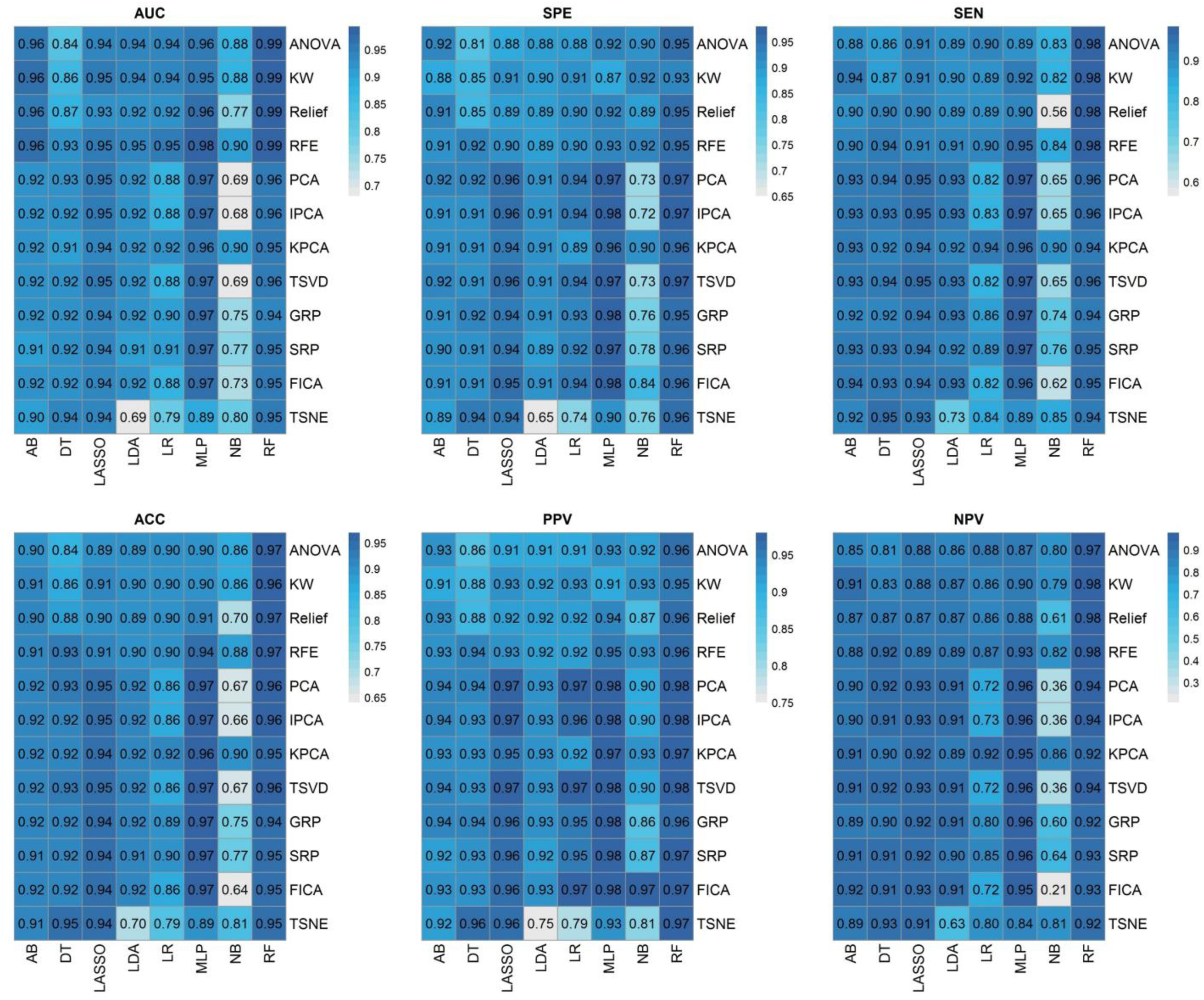
Heatmaps of the cross-combination of feature selectors (12 rows) and classifiers (8 columns) for AUC, sensitivity, specificity, accuracy, PPV, NPV in dataset #2.

For dataset #3, Figure 6 represents the heatmap of the cross-combination of feature selectors and classifiers for AUC, sensitivity, specificity, accuracy, PPV, NPV, which CT images with PCR-Positive and community-acquired pneumonia were included and then randomly split into 70% (10,205 patients) and 30% (4,200 patients) for the training and test sets, respectively. ANOVA feature selection and RF classifier combination resulted in highest performance (AUC = 0.98, sensitivity = 0.96, specificity = 93, accuracy = 0.94, PPV = 0.93, NPV = 0.96). Supplemental Figure 2 represents a box plot of feature selectors and classifiers for different evaluation metrics in dataset #3. Figure 7 represents the receiver operating characteristic curve for three different strategies. Supplemental figures 8-10 represent the statistical comparison of AUC between the different models using DeLong test.

**Figure 6.**
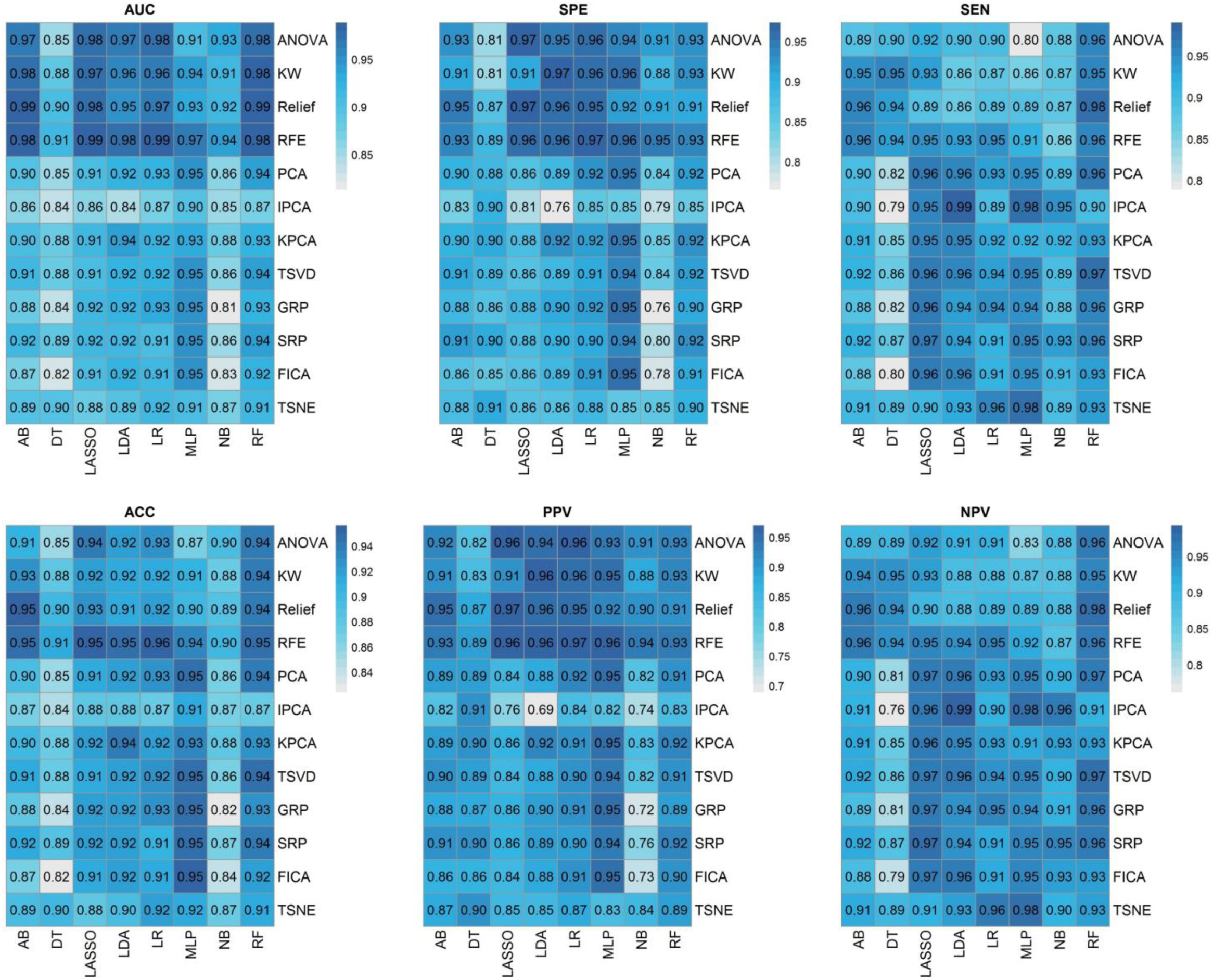
Heatmaps of the cross-combination of feature selectors (12 rows) and classifiers (8 columns) for AUC, sensitivity, specificity, accuracy, PPV, NPV in dataset #3.

**Figure 7.**
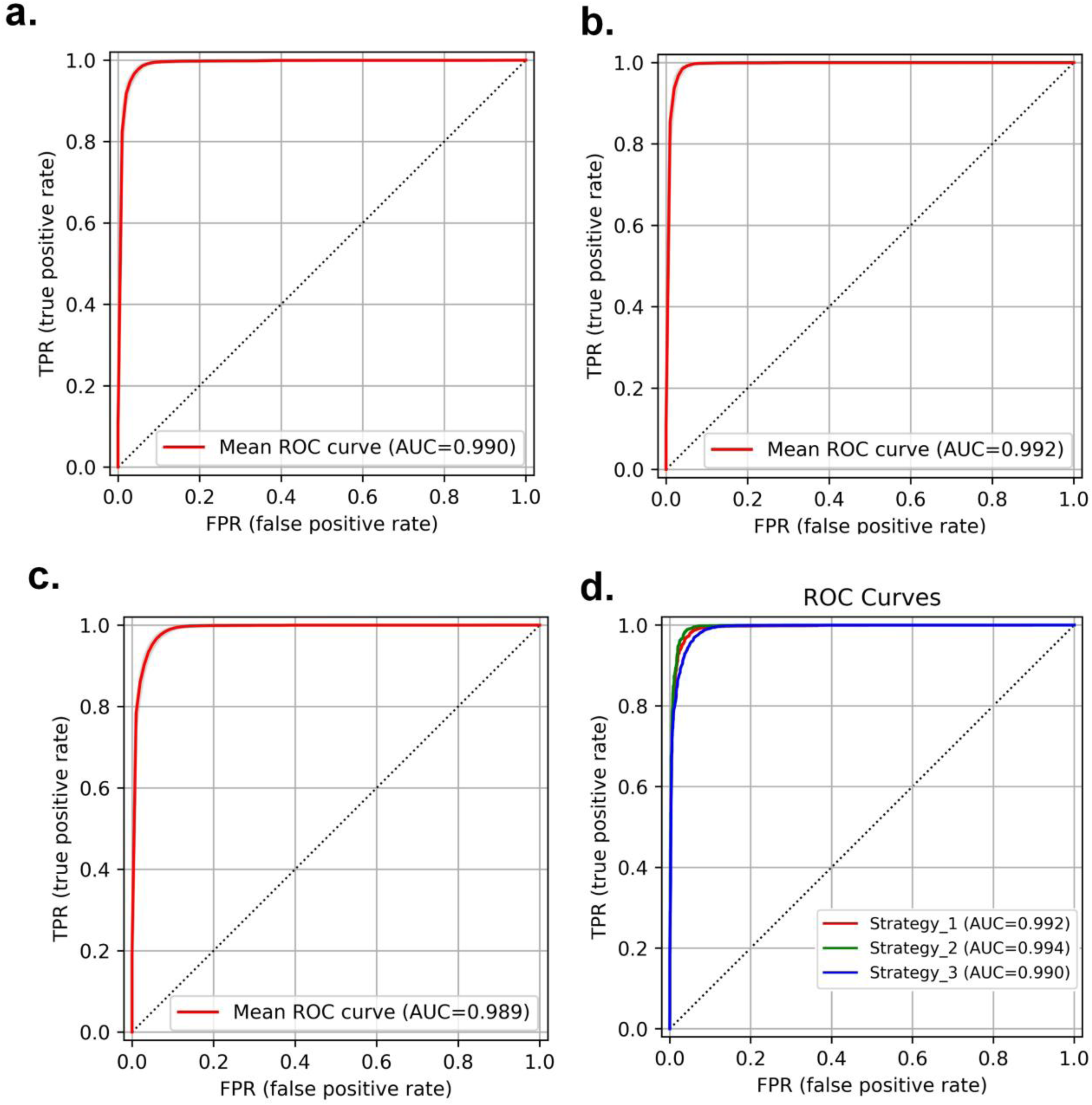
ROC curves for dataset #1 (a), dataset #2 (b) and dataset #3 (c). For figure (a, b and c), confidence intervals are also plotted (gray) with 10000 bootstrapping. The confidence intervals are very small due to the large scale of our study. In figure (d), the results from strategies 1-3 are plotted all on one figure to compare the different algorithms.

## DISCUSSION

In this work, we implemented multiple machine learning algorithms to detect COVID-19 in a dataset consisting of a large number of patients (COVID-19, non-COVID-19 pneumonia, lung cancer, PE and normal patients). The entire lung was segmented using automated DL algorithms, and subsequently radiomics features were extracted. Features were normalized, redundant features eliminated, and remaining features fed to feature selection algorithms and classifiers. Our models were evaluated using different strategies. We successfully discriminated COVID-19 from other lung diseases and also COVID-19 pneumonia from other pneumonia.

Our findings are consistent with previous studies focusing on the same goals. A study by Fang et al.^54^ showed that a radiomics model based on CT imaging features could differentiate COVID-19 pneumonia from other types of pneumonia with an area under the ROC curve of 95%. Their results also suggested that radiomics perform better than the clinical-only model. In another study, Tan et al. ^55^ demonstrated the efficacy of a radiomics-based model in discovering whether a patient has COVID-19 pneumonia or other types of pneumonia by analyzing the non-infectious areas of their CT scan. Their model achieved an AUC as high as 0.95 both in the training and test datasets. Di et al. ^37^ also studied the diagnostic accuracy of CT-based radiomics in patients and reached a robust model. Their hypergraph model could distinguish between community-acquired pneumonia and COVID-19 disease pneumonia.

Although some studies investigated the power of radiomics in the differentiation of lung abnormalities as mentioned earlier. Several research studies were conducted on the application of other AI methods, such as DL/ML algorithms, and DL + radiomics models. A study by Yousefzadeh et al. ^56^ investigated the performance of a DL-based model (ai-corona) in the differentiation task between COVID-19 pneumonia, other pneumonia, non-pneumonia, and normal images. Their model reached an AUC as high as 0.997, 0.989, and 0.954 in three different test sets. Harmon et al. ^34^ trained a DL algorithm on a multi-national dataset of 1,280 patients and evaluated its classification ability using a chest CT dataset of 1,337 patients. They achieved an accuracy of 90.8%. Ni et al. ^57^ also studied a DL model aiming to locate and detect lung abnormalities, including COVID-19 pneumonia. Their model reached 0.97 in the F1 score and assisted the radiologists in faster detection and diagnosis (p-values<0.0001).

Similar to most radiomics studies of COVID-19, we used chest CT images ^37, 38, 58^. However, there is ongoing research exploiting other imaging modalities as well. For example, Bae et al. ^59^ evaluated the prediction ability of radiomics in chest X-rays of 514 patients. They attempted to gain assistance from other methods DL (Radiomics feature Map + DL + clinical features) and achieved AUCs of 0.93 and 0.90 in the prediction of mortality and the need for mechanical ventilators, respectively. In another study by Chandra et al. ^60^, chest X-rays of 2,088 (training set) and 258 (testing set) patients taken at baseline, were assessed and radiomics analysis performed. They reached an AUC of 0.95 in the test set for identifying normal, suspicious, and COVID-19 groups of patients.

On the use of different lung pathologies in the dataset, several other studies have been conducted. The dataset of Amyar et al. ^61^ covered healthy patients as well as those with COVID-19, lung cancer, and some other lung pathologies. They achieved an AUC of 0.97 for the classification task. Wang et al. ^41^ utilized a PARL (prior attention residual learning)-based model for the classification of CT images into COVID-19 pneumonia, other pneumonia, and non-pneumonic images and could achieve a AUC of 0.97 for COVID-19 discrimination. Chen et al. ^62^ included 422 patients who had COVID-19, other types of pneumonia, tuberculosis, and also normal images in their dataset. Their ResNet model achieved an accuracy of 91.2% for classifying images. Our study also included several pathologies in the lung, including COVID-19, pneumonia of other types (viral and bacterial), lung cancer, and also normal lung images.

Overall, several studies assessed radiomics and DL and DL + Radiomics algorithms for diagnostic and classification purposes with a small size dataset in most cases ^36, 63, 64^. In this work, we concluded that radiomic features combined with appropriate machine learning algorithm has the potential to enhance our diagnostic ability in the differentiation of COVID-19 pneumonia, other types of pneumonia, and several other lung diseases. We utilized a large-sized dataset consisting of CT images of patients from multiple institutions (multiple scanners/ imaging protocols) from different countries. The large-scale dataset of our study helped the generalizability and reproducibility of our model, which was evaluated using different scenarios.

Our efforts faced a number of limitations, some of which were addressed and considered. For example, motion artefacts are inescapable when patients are undergoing CT scans. Hence, we excluded these patients as they had an overlapping region of pneumonia in their chest image. Second, not all patients were tested for COVID-19 RT-PCR as some of them were included in our study only by their positive CT signs. Thus, we attempted to overcome this limitation using different model evaluation scenarios (e.g. choosing patients with positive RT-PCR as the testing set) in order to reach a reproducible model for further studies. Third, we did not include clinical or laboratory data. However, these data have been shown to be linked with CT image features ^36, 65^. Finally, image acquisition parameters were undoubtedly distinct in each center and affected radiomic features. Further studies should be performed using harmonized features between different centers.

## CONCLUSION

We evaluated multiple machine learning algorithms to detect COVID-19 pneumonia and discriminate it from other lung diseases using radiomics features of the entire lung using a very large heterogeneous dataset. We successfully discriminated COVID-19 from other lung diseases and also COVID-19 from other community-acquired pneumonia. Radiomic features of whole lung and machine learning algorithms combination could effectively detect COVID-19 cases to boost clinical diagnosis without the need for other diagnostic tests.

## Data and code availability

Radiomics features and code will be available upon request.

## Data Availability

All data produced in the present work are contained in the manuscript

https://www.cancerimagingarchive.net/collections/

https://mosmed.ai/datasets/covid19_1110

http://ictcf.biocuckoo.cn/HUST-19.php

https://www.rsna.org/education/ai-resources-and-training/ai-image-challenge

## Acknowledgements

This work was supported by the Swiss National Science Foundation under grant SNRF 320030_176052.

---

| Section/Topic |  | Checklist Item |  | Page |
| --- | --- | --- | --- | --- |
| Title and abstract |  |  |  |  |
| Title | 1 | D;V | Identify the study as developing and/or validating a multivariable prediction model, the target population, and the outcome to be predicted. | 1 |
| Abstract | 2 | D;V | Provide a summary of objectives, study design, setting, participants, sample size, predictors, outcome, statistical analysis, results, and conclusions. | 1 |
| Introduction |  |  |  |  |
| Background and objectives | 3a | D;V | Explain the medical context (including whether diagnostic or prognostic) and rationale for developing or validating the multivariable prediction model, including references to existing models. | 2-3 |
|  | 3b | D;V | Specify the objectives, including whether the study describes the development or validation of the model or both. | 3 |
| Methods |  |  |  |  |
| Source of data | 4a | D;V | Describe the study design or source of data (e.g., randomized trial, cohort, or registry data), separately for the development and validation data sets, if applicable. | 4 |
|  | 4b | D;V | Specify the key study dates, including start of accrual; end of accrual; and, if applicable, end of follow-up. | 4 |
| Participants | 5a | D;V | Specify key elements of the study setting (e.g., primary care, secondary care, general population) including number and location of centres. | 4-5 |
|  | 5b | D;V | Describe eligibility criteria for participants. | 4 |
|  | 5c | D;V | Give details of treatments received, if relevant. | NA |
| Outcome | 6a | D;V | Clearly define the outcome that is predicted by the prediction model, including how and when assessed. | 4,6 |
|  | 6b | D;V | Report any actions to blind assessment of the outcome to be predicted. | 5-6 |
| Predictors | 7a | D;V | Clearly define all predictors used in developing or validating the multivariable prediction model, including how and when they were measured. | 5-6 |
|  | 7b | D;V | Report any actions to blind assessment of predictors for the outcome and other predictors. | 5-6 |
| Sample size | 8 | D;V | Explain how the study size was arrived at. | 4 |
| Missing data | 9 | D;V | Describe how missing data were handled (e.g., complete-case analysis, single imputation, multiple imputation) with details of any imputation method. | NA |
| Statistical analysis methods | 10a | D | Describe how predictors were handled in the analyses. | 5 |
|  | 10b | D | Specify type of model, all model-building procedures (including any predictor selection), and method for internal validation. | 5 |
|  | 10c | V | For validation, describe how the predictions were calculated. | 5-6 |
|  | 10d | D;V | Specify all measures used to assess model performance and, if relevant, to compare multiple models. | 5-6 |
|  | 10e | V | Describe any model updating (e.g., recalibration) arising from the validation, if done. | 5-6 |
| Risk groups | 11 | D;V | Provide details on how risk groups were created, if done. | NA |
| Development vs. validation | 12 | V | For validation, identify any differences from the development data in setting, eligibility criteria, outcome, and predictors. | 5-6 |
| Results |  |  |  |  |
| Participants | 13a | D;V | Describe the flow of participants through the study, including the number of participants with and without the outcome and, if applicable, a summary of the follow-up time. A diagram may be helpful. | 5 |
|  | 13b | D;V | Describe the characteristics of the participants (basic demographics, clinical features, available predictors), including the number of participants with missing data for predictors and outcome. Not mentioned in result | 5 |
|  | 13c | V | For validation, show a comparison with the development data of the distribution of important variables (demographics, predictors and outcome). | 6-7 |
| Model development | 14a | D | Specify the number of participants and outcome events in each analysis. | 5 |
|  | 14b | D | If done, report the unadjusted association between each candidate predictor and outcome. | 6-7 |
| Model specification | 15a | D | Present the full prediction model to allow predictions for individuals (i.e., all regression coefficients, and model intercept or baseline survival at a given time point). | 6-7 |
|  | 15b | D | Explain how to the use the prediction model. | 6-7 |
| Model performance | 16 | D;V | Report performance measures for the prediction model. | 6-7 |
| Model-updating | 17 | V | If done, report the results from any model updating (i.e., model specification, model performance). | 6-7 |
| Discussion |  |  |  |  |
| Limitations | 18 | D;V | Discuss any limitations of the study (such as nonrepresentative sample, few events per predictor, missing data). | 9 |
| Interpretation | 19a | V | For validation, discuss the results with reference to performance in the development data, and any other validation data. | 7-8 |
|  | 19b | D;V | Give an overall interpretation of the results, considering objectives, limitations, results from similar studies, and other relevant evidence. | 7-9 |
| Implications | 20 | D;V | Discuss the potential clinical use of the model and implications for future research. | 7-9 |
| Other information |  |  |  |  |
| Supplementary information | 21 | D;V | Provide information about the availability of supplementary resources, such as study protocol, Web calculator, and data sets. | 9 |
| Funding | 22 | D;V | Give the source of funding and the role of the funders for the present study. | 9 |

\*Items relevant only to the development of a prediction model are denoted by D, items relating solely to a validation of a prediction model are denoted by V, and items relating to both are denoted D;V. We recommend using the TRIPOD Checklist in conjunction with the TRIPOD Explanation and Elaboration document.

CLAIM: Checklist for Artificial Intelligence in Medical Imaging
| Section / Topic | No. | Item |  |
| --- | --- | --- | --- |
| **TITLE / ABSTRACT** |  |  |  |
|  | **1** | Identification as a study of AI methodology, specifying the category of technology used (e.g., deep learning) | ✓ |
|  | **2** | Structured summary of study design, methods, results, and conclusions | ✓ |
| **INTRODUCTION** |  |  |  |
|  | **3** | Scientific and clinical background, including the intended use and clinical role of the AI approach | ✓ |
|  | **4** | Study objectives and hypotheses | ✓ |
| **METHODS** |  |  |  |
| *Study Design* | **5** | Prospective or retrospective study | ✓ |
|  | **6** | Study goal, such as model creation, exploratory study, feasibility study, non-inferiority trial | ✓ |
| *Data* | **7** | Data sources | ✓ |
|  | **8** | Eligibility criteria: how, where, and when potentially eligible participants or studies were identified (e.g., symptoms, results from previous tests, inclusion in registry, patient-care setting, location, dates) | ✓ |
|  | **9** | Data pre-processing steps | ✓ |
|  | **10** | Selection of data subsets, if applicable | ✓ |
|  | **11** | Definitions of data elements, with references to Common Data Elements | ✓ |
|  | **12** | De-identification methods | ✓ |
|  | **13** | How missing data were handled | ✓ |
| *Ground Truth* | **14** | Definition of ground truth reference standard, in sufficient detail to allow replication | ✓ |
|  | **15** | Rationale for choosing the reference standard (if alternatives exist) | ✓ |
|  | **16** | Source of ground-truth annotations; qualifications and preparation of annotators | ✓ |
|  | **17** | Annotation tools | ✓ |
|  | **18** | Measurement of inter- and intrarater variability; methods to mitigate variability and/or resolve discrepancies | - |
| *Data Partitions* | **19** | Intended sample size and how it was determined | - |
|  | **20** | How data were assigned to partitions; specify proportions | ✓ |
|  | **21** | Level at which partitions are disjoint (e.g., image, study, patient, institution) | ✓ |
| *Model* | **22** | Detailed description of model, including inputs, outputs, all intermediate layers and connections | ✓ |
|  | **23** | Software libraries, frameworks, and packages | ✓ |
|  | **24** | Initialization of model parameters (e.g., randomization, transfer learning) | ✓ |
| *Training* | **25** | Details of training approach, including data augmentation, hyperparameters, number of models trained | ✓ |
|  | **26** | Method of selecting the final model | ✓ |
|  | **27** | Ensembling techniques, if applicable | ✓ |
| *Evaluation* | **28** | Metrics of model performance | ✓ |
|  | **29** | Statistical measures of significance and uncertainty (e.g., confidence intervals) | ✓ |
|  | **30** | Robustness or sensitivity analysis | - |
|  | **31** | Methods for explainability or interpretability (e.g., saliency maps), and how they were validated | - |
|  | **32** | Validation or testing on external data | ✓ |
| **RESULTS** |  |  |  |
| *Data* | **33** | Flow of participants or cases, using a diagram to indicate inclusion and exclusion | - |
|  | **34** | Demographic and clinical characteristics of cases in each partition | ✓ |
| *Model performance* | **35** | Performance metrics for optimal model(s) on all data partitions | ✓ |
|  | **36** | Estimates of diagnostic accuracy and their precision (such as 95% confidence intervals) | ✓ |
|  | **37** | Failure analysis of incorrectly classified cases | - |
| **DISCUSSION** |  |  |  |
|  | **38** | Study limitations, including potential bias, statistical uncertainty, and generalizability | ✓ |
|  | **39** | Implications for practice, including the intended use and/or clinical role | ✓ |
| **OTHER INFORMATION** |  |  |  |
|  | **40** | Registration number and name of registry | - |
|  | **41** | Where the full study protocol can be accessed | - |
|  | **42** | Sources of funding and other support; role of funders | ✓ |

**Figure 1:**
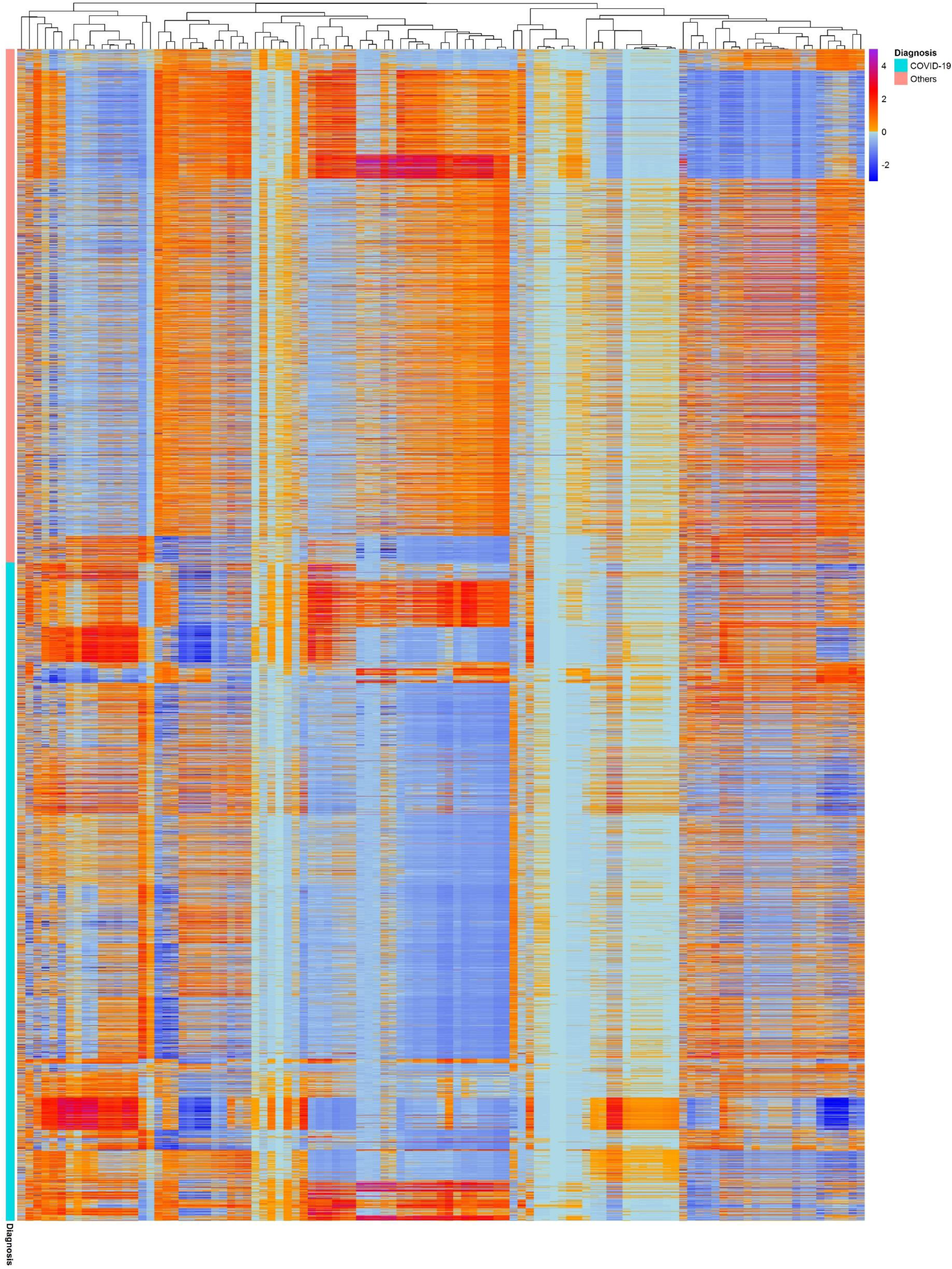
Cluster heat map of dataset 2, columns and rows are the radiomics features and patients sample, respectively

**Figure 2:**
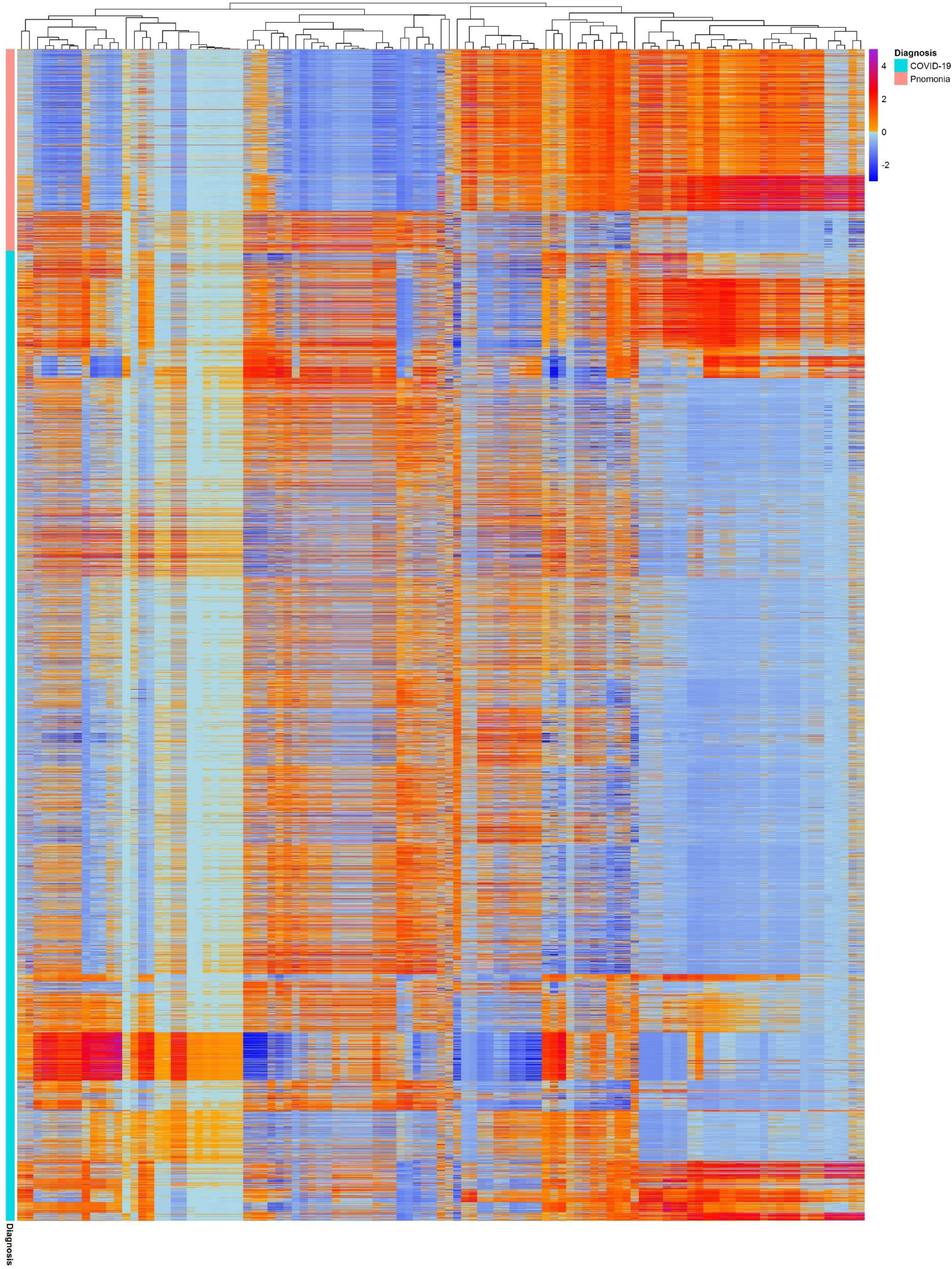
Cluster heat map for dataset 3. columns and rows are the radiomics features and patients sample, respectively

**Figure 3:**
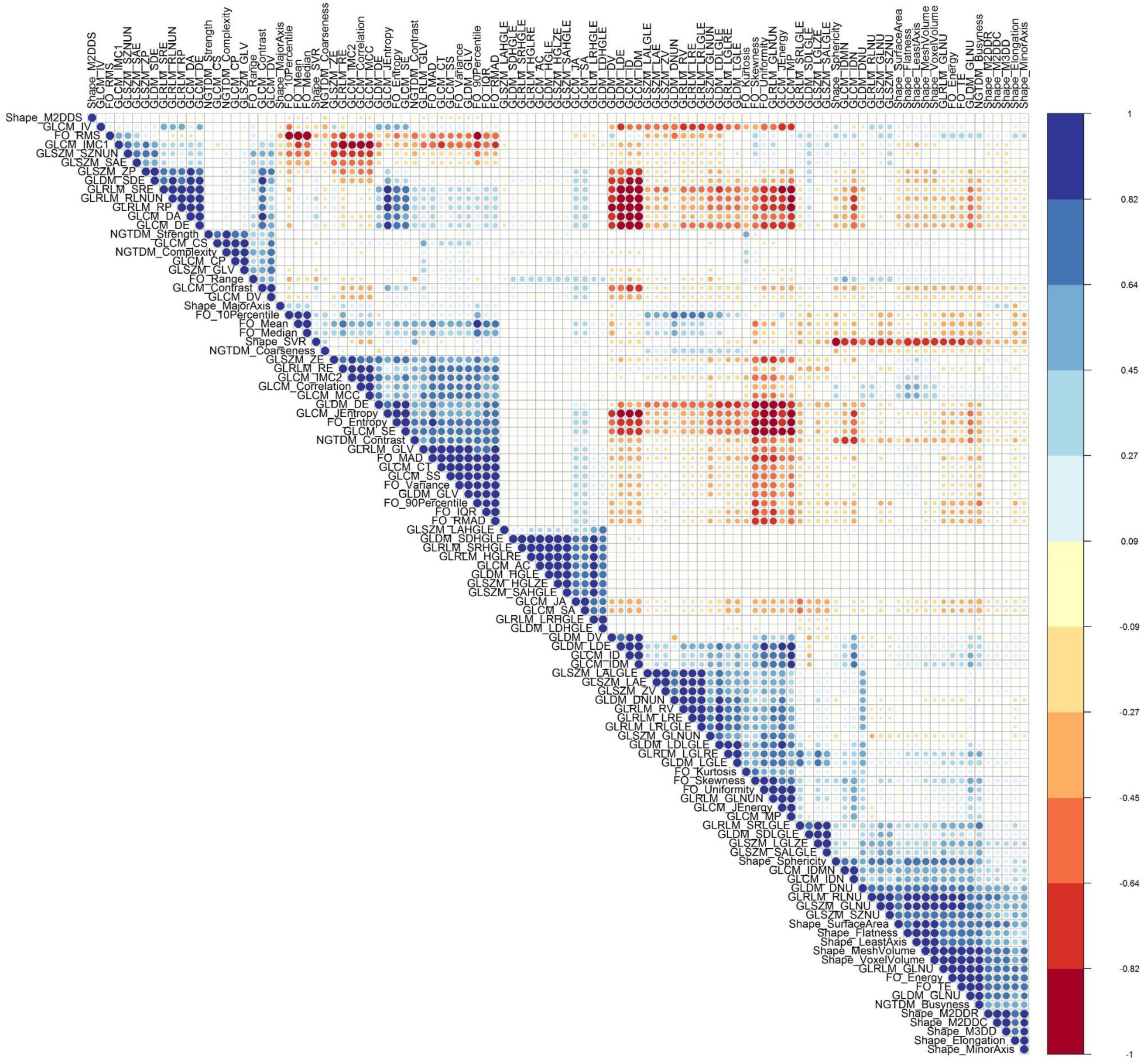
Correlation matrix between different features in dataset 2

**Figure 4:**
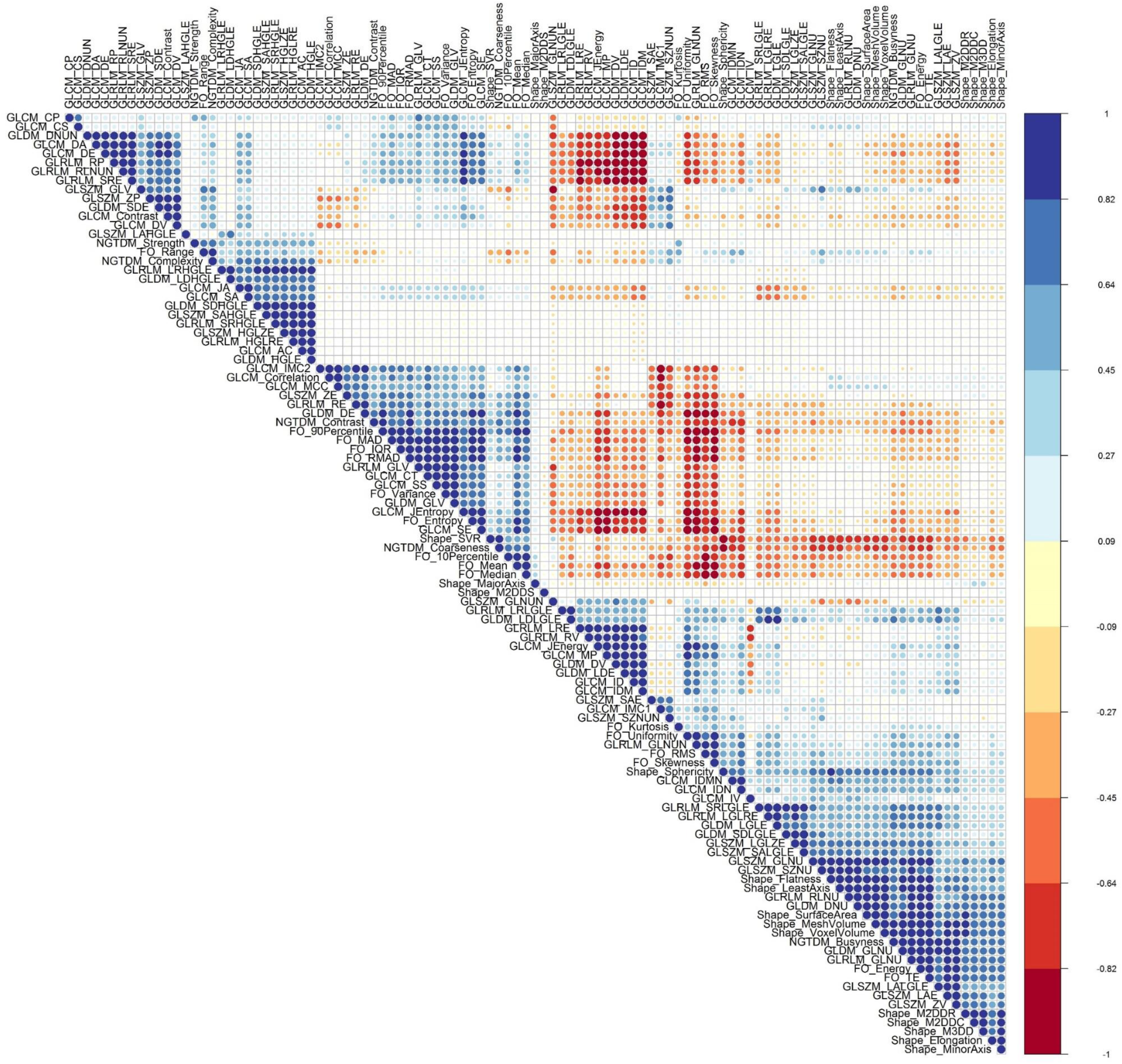
Correlation matrix between different features in dataset 3

**Figure 5:**
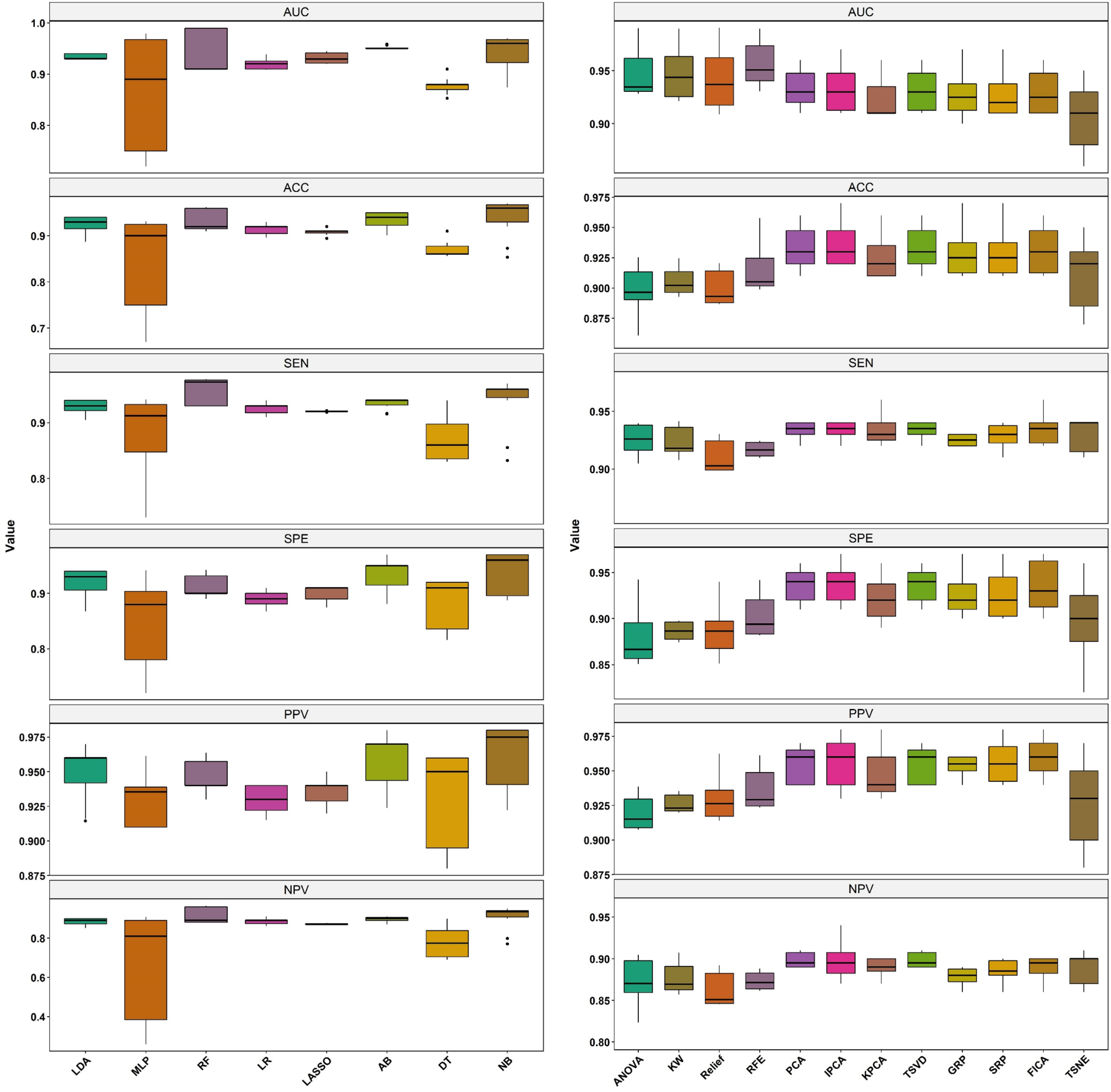
Box plot of different quantitative parameters for different feature selectors and classifiers in dataset 1

**Figure 6:**
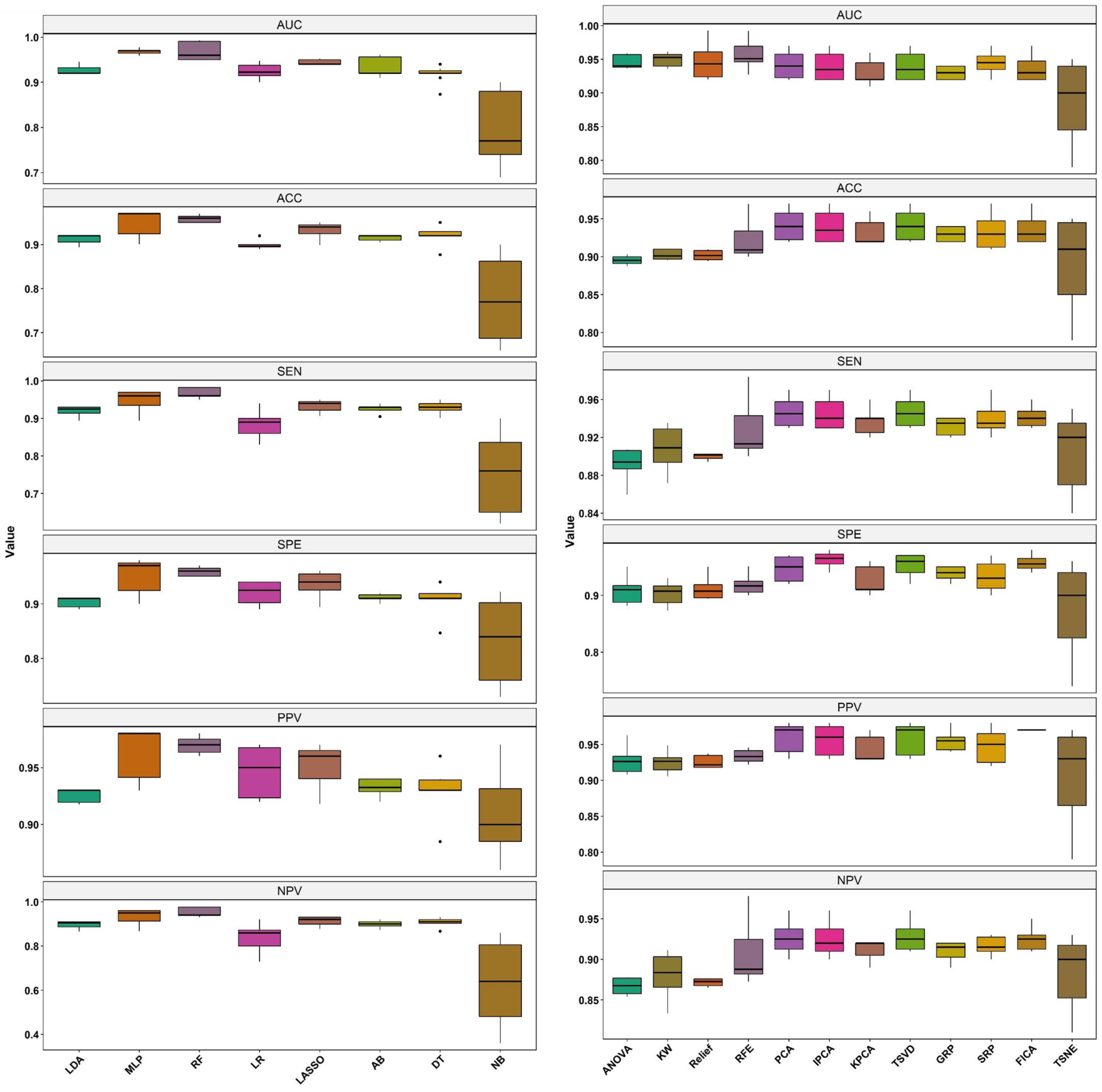
Box plot of different quantitative parameters for different feature selectors and classifiers for dataset 2

**Figure 7:**
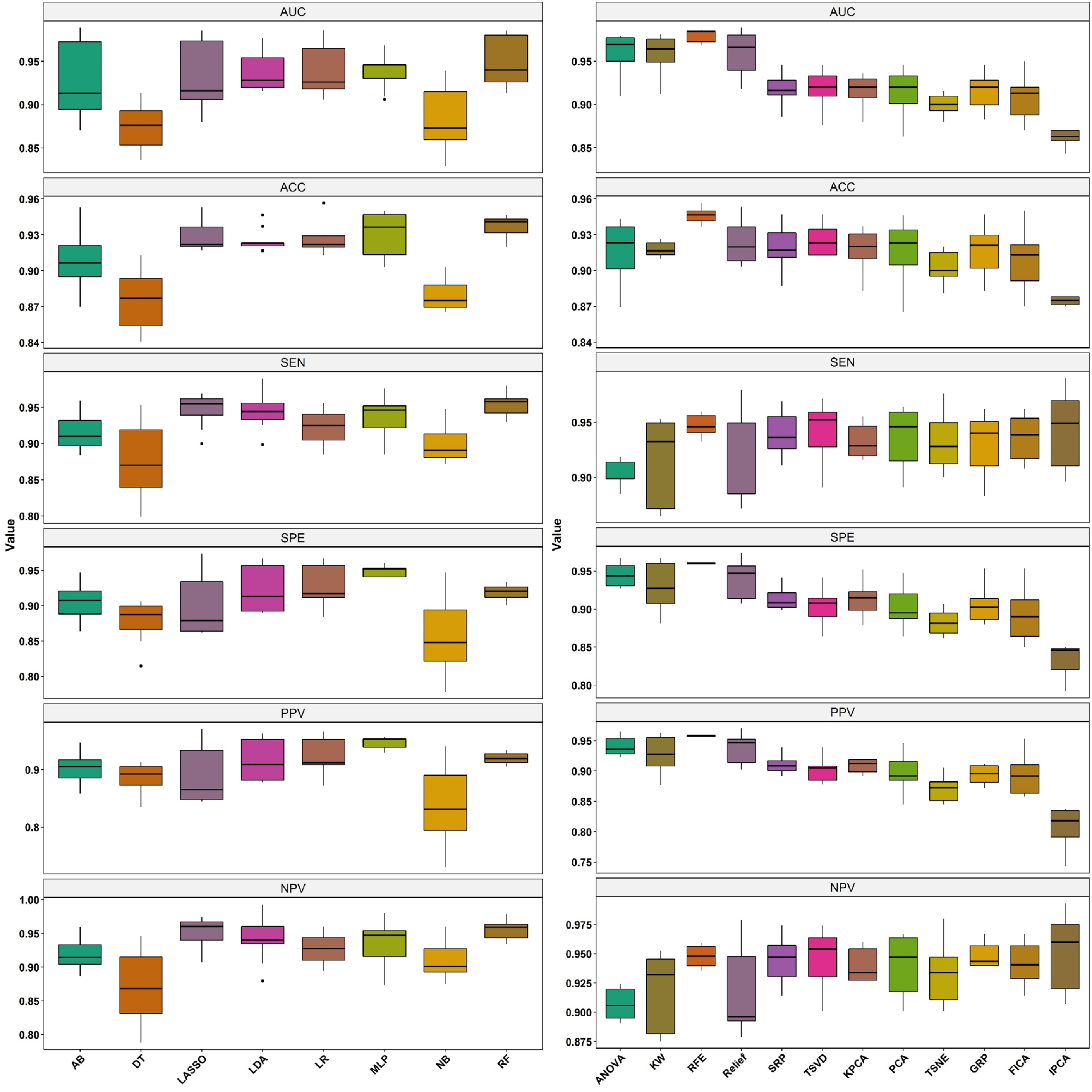
Box plot of different quantitative parameters for different feature selectors and classifiers for dataset 3

**Figure 8:**
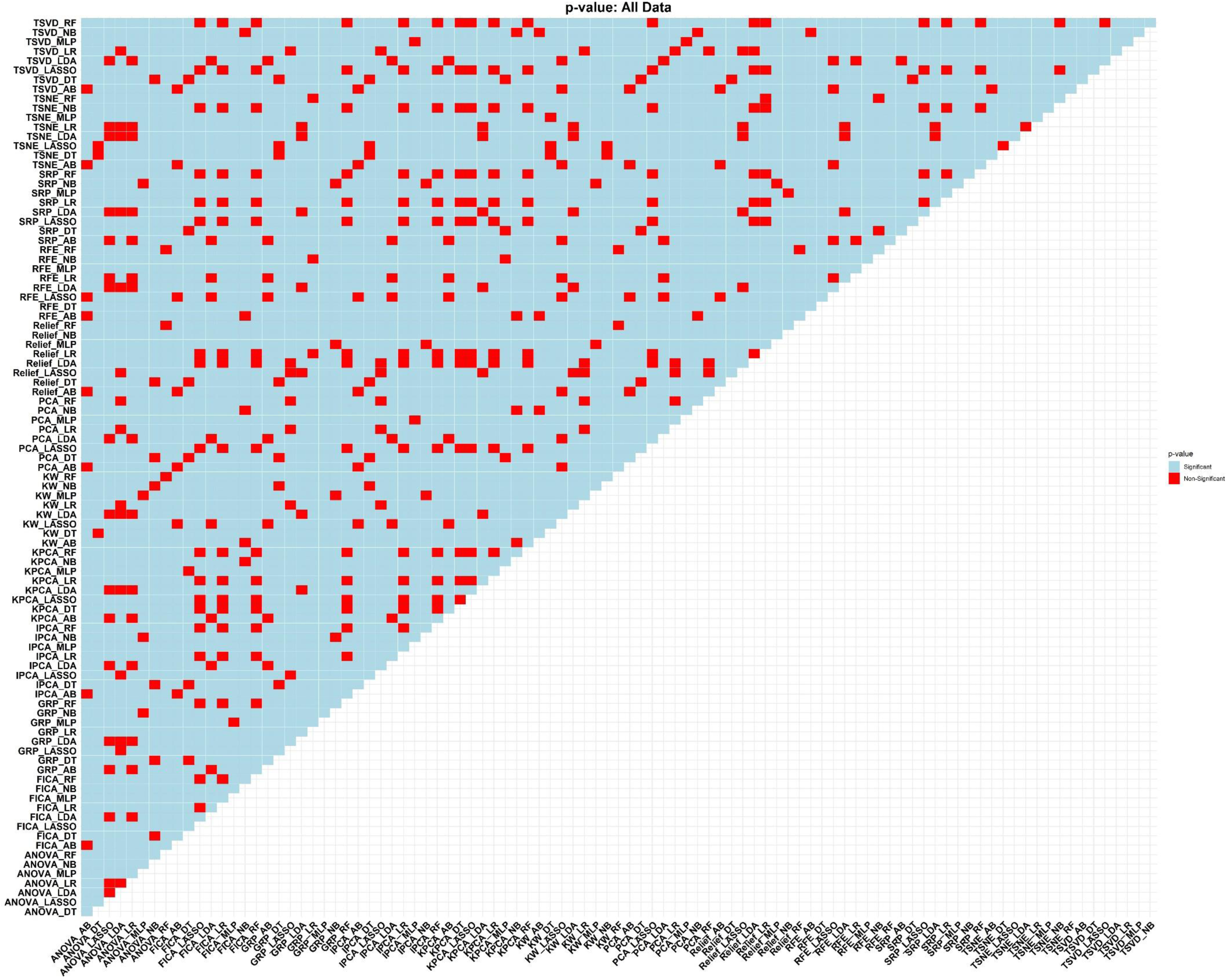
Statistical comparison of AUC values for different model using DeLong test in dataset 1

**Figure 9:**
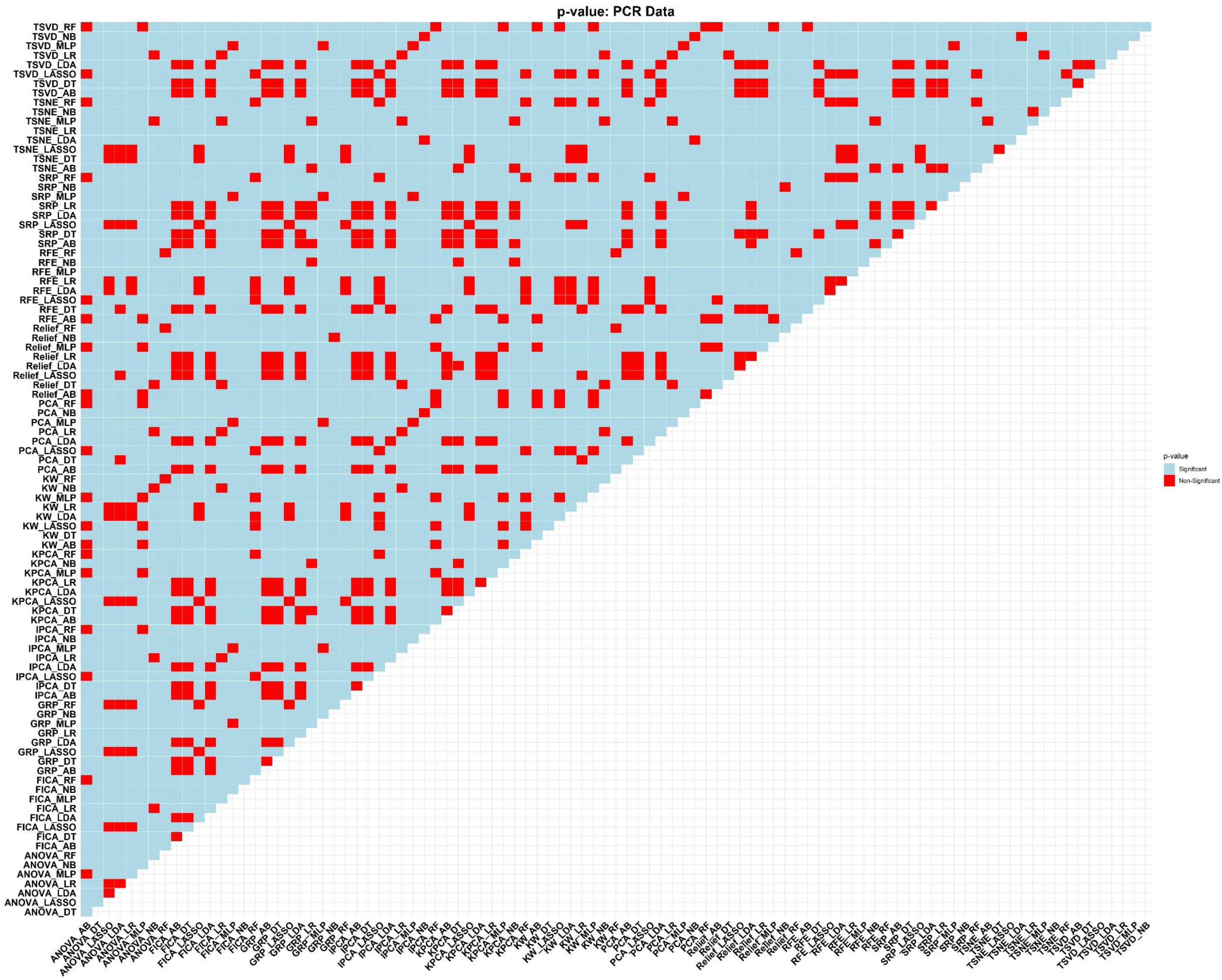
Statistical comparison of AUC values for different models using DeLong test in dataset 2

**Figure 10:**
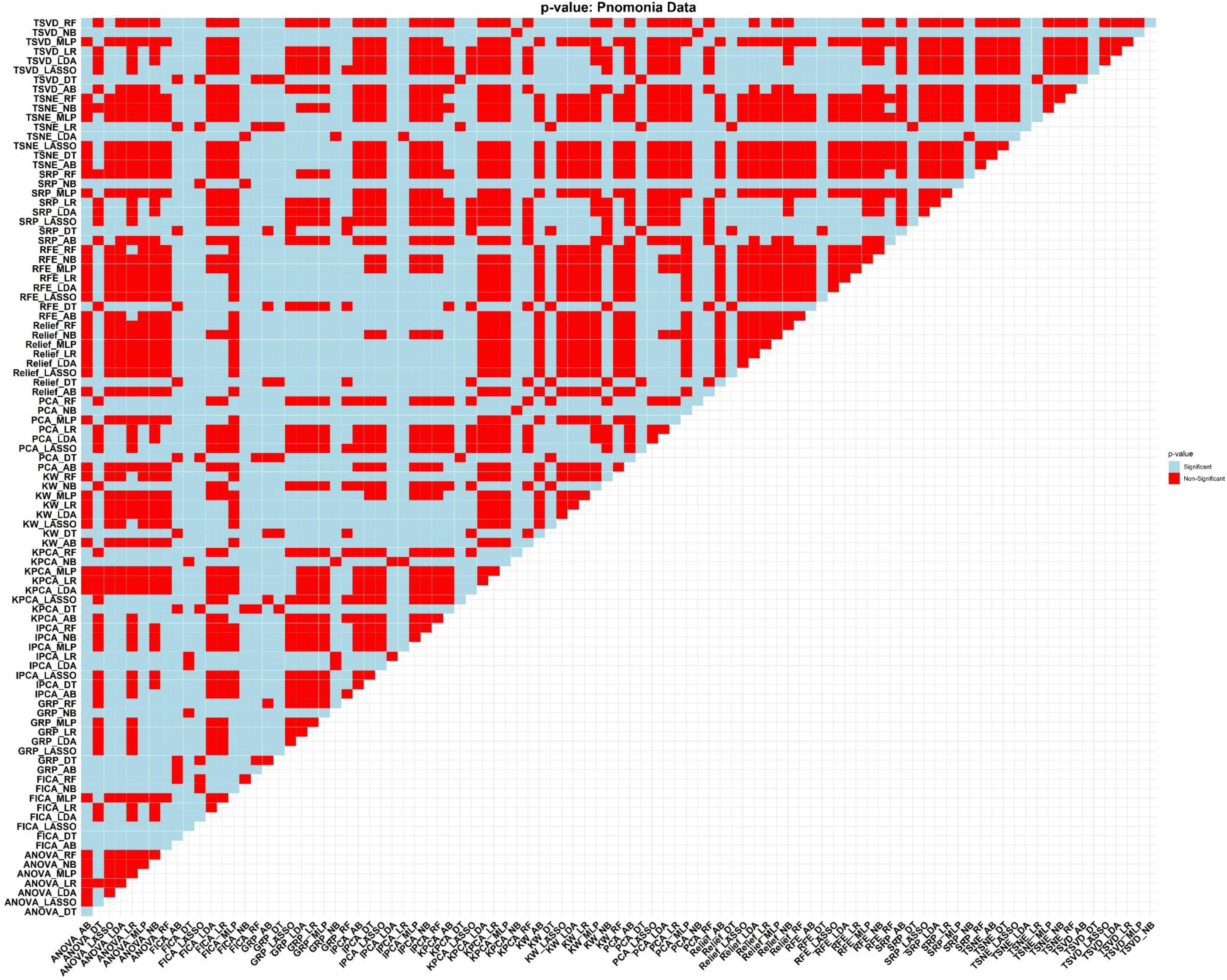
Statistical comparison of AUC values for different models using DeLong test in dataset 3

**Table 1:**
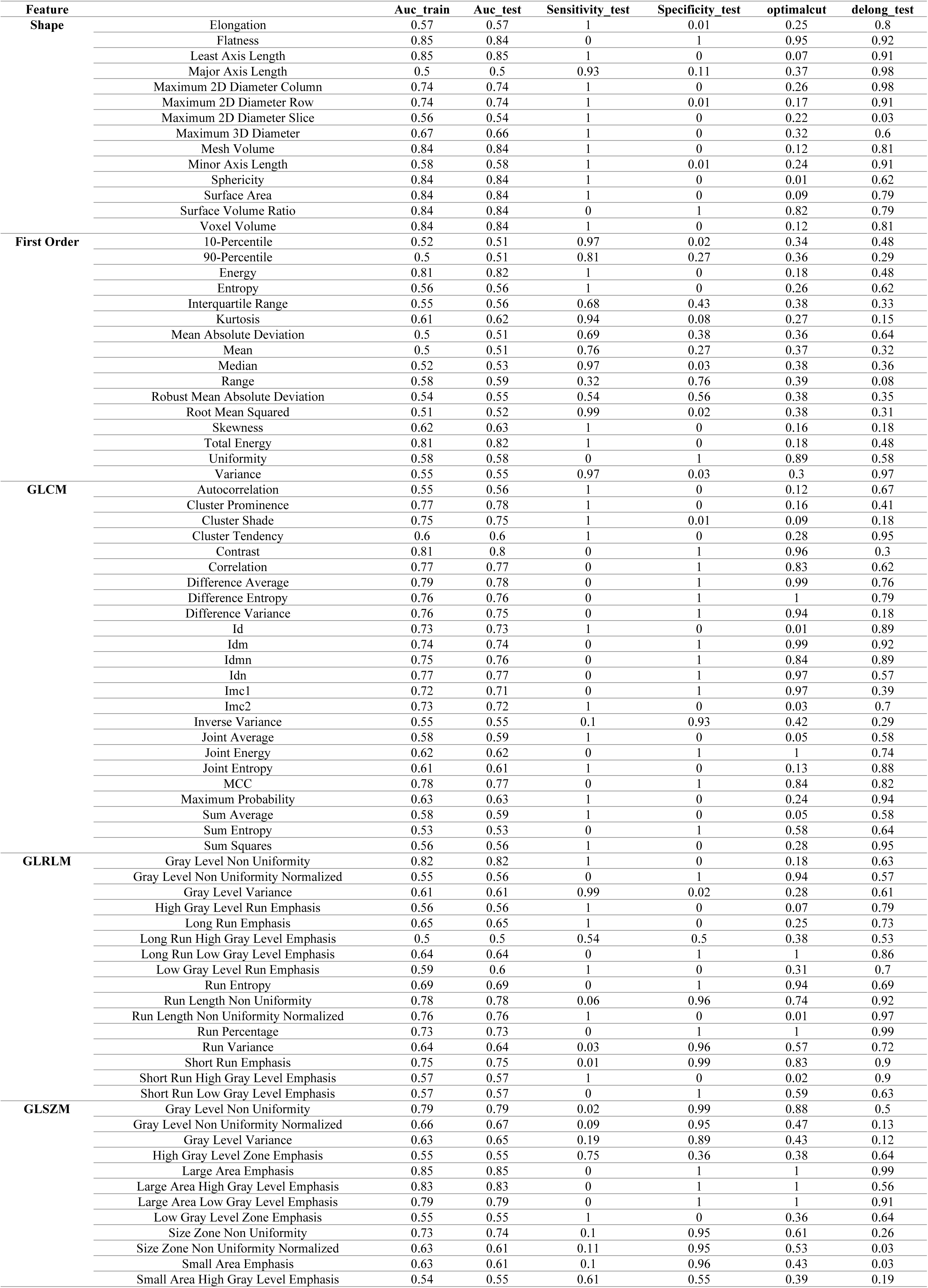
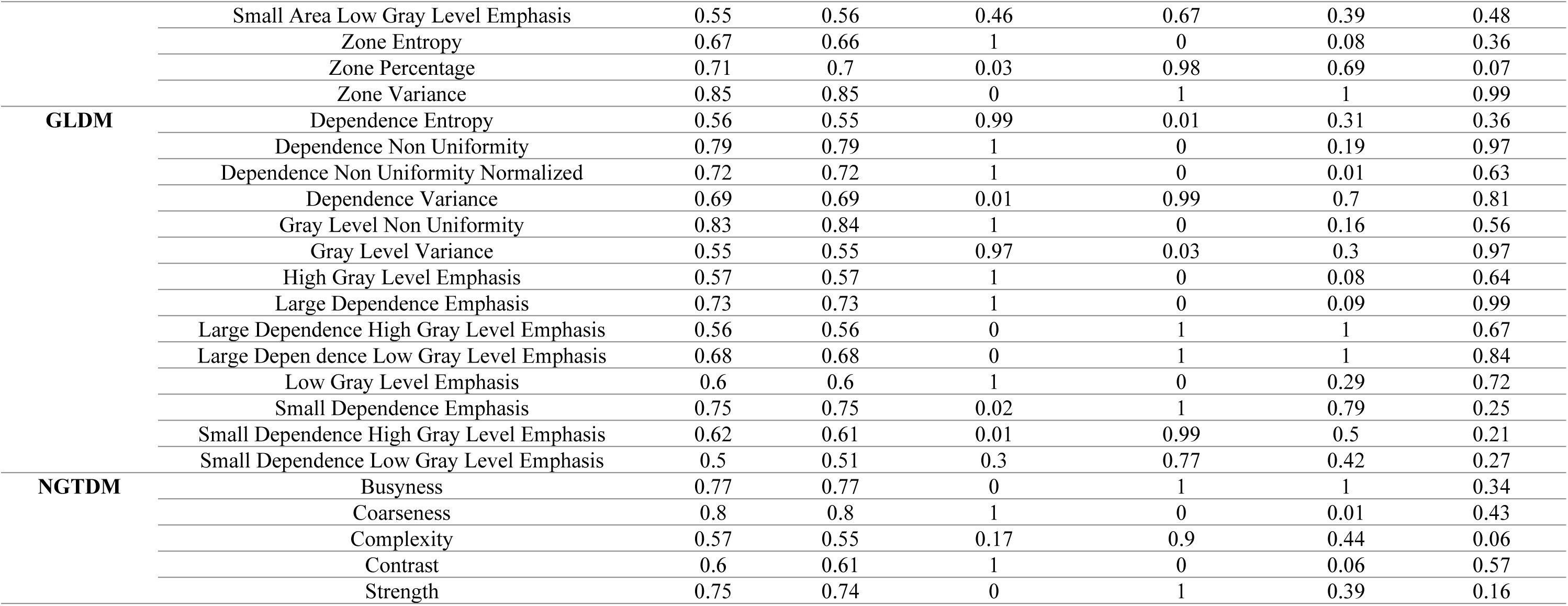
Univariate analysis in dataset 1

**Table 2:**
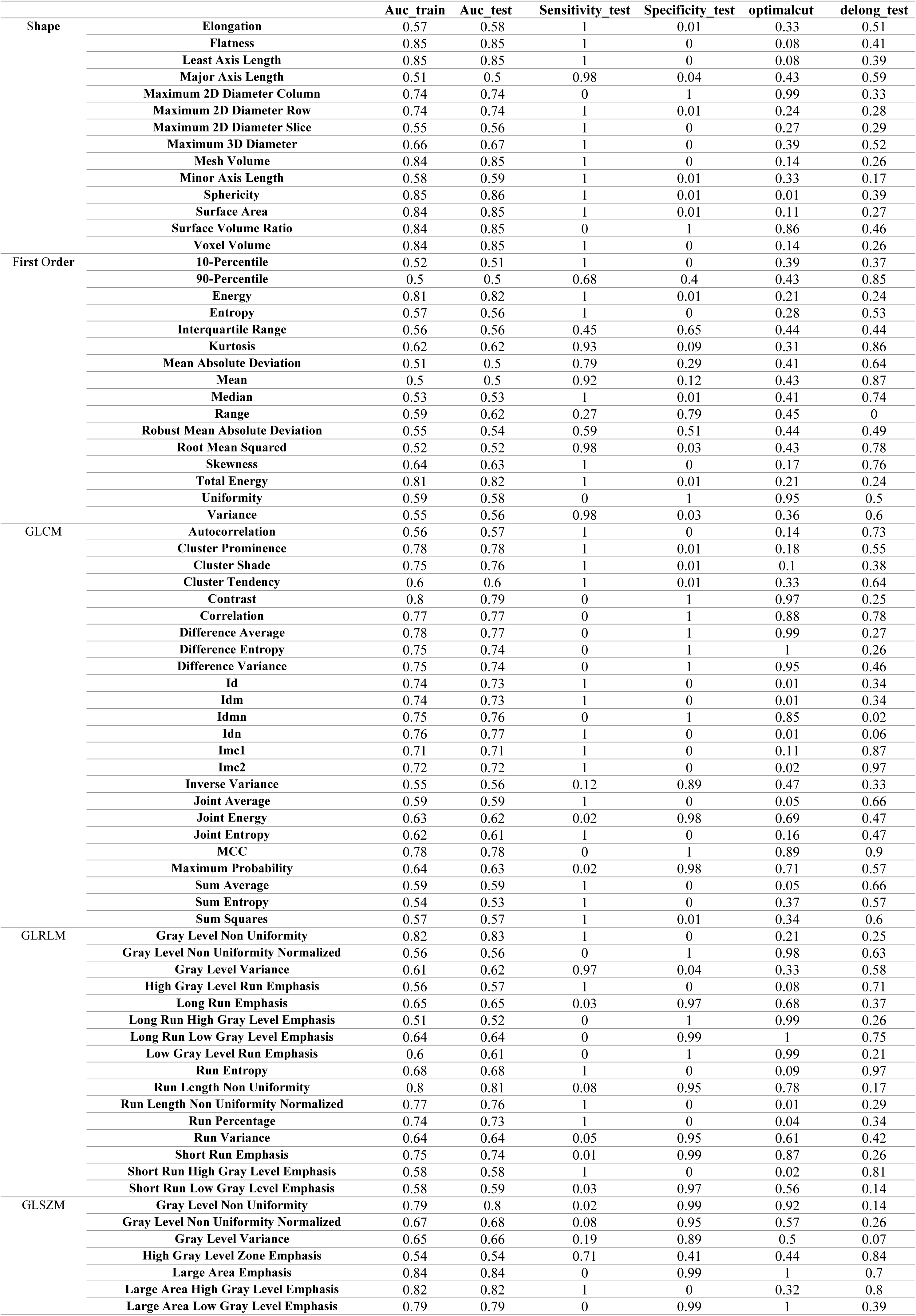
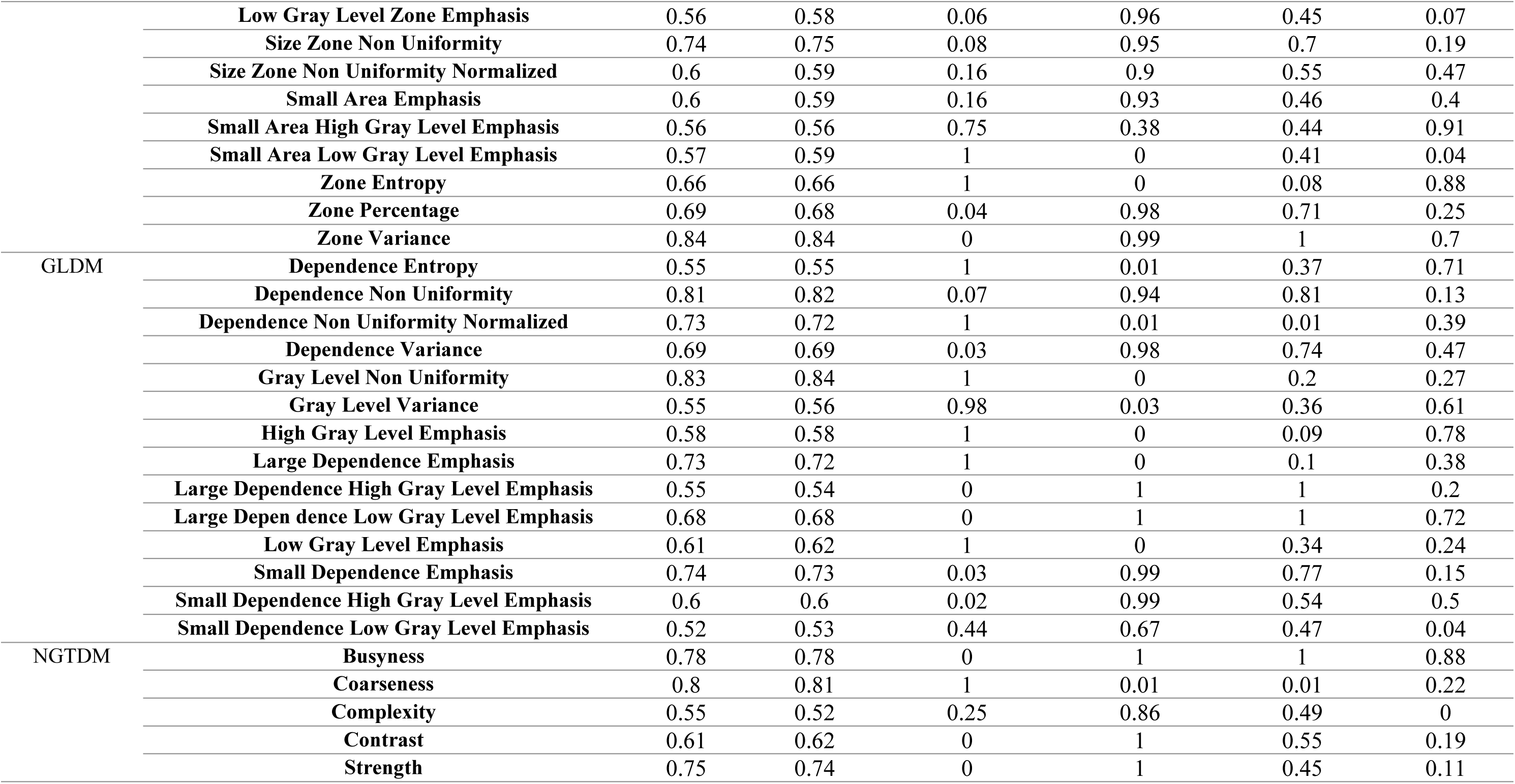
Univariate analysis in dataset 2

**Table 3:**
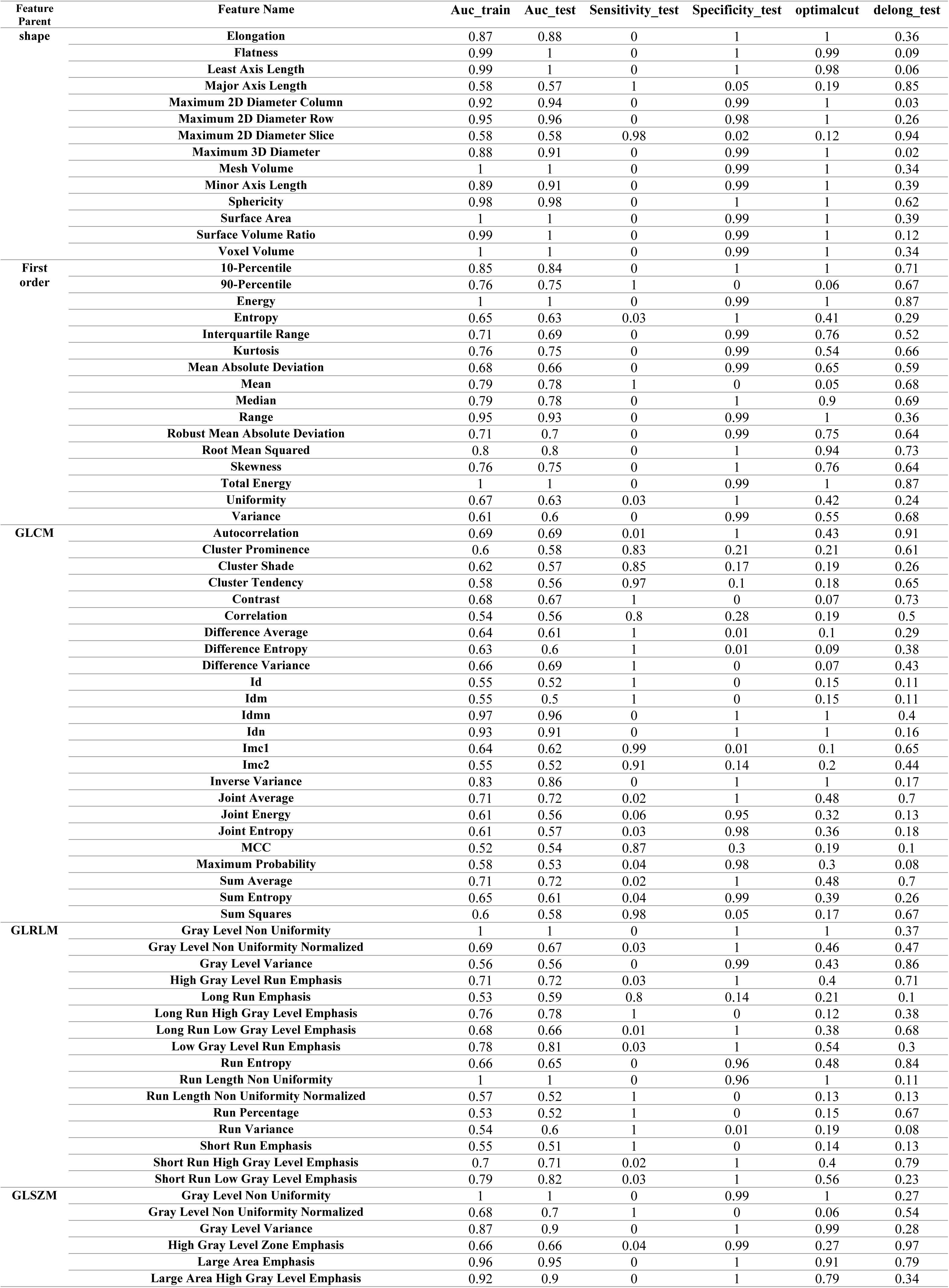
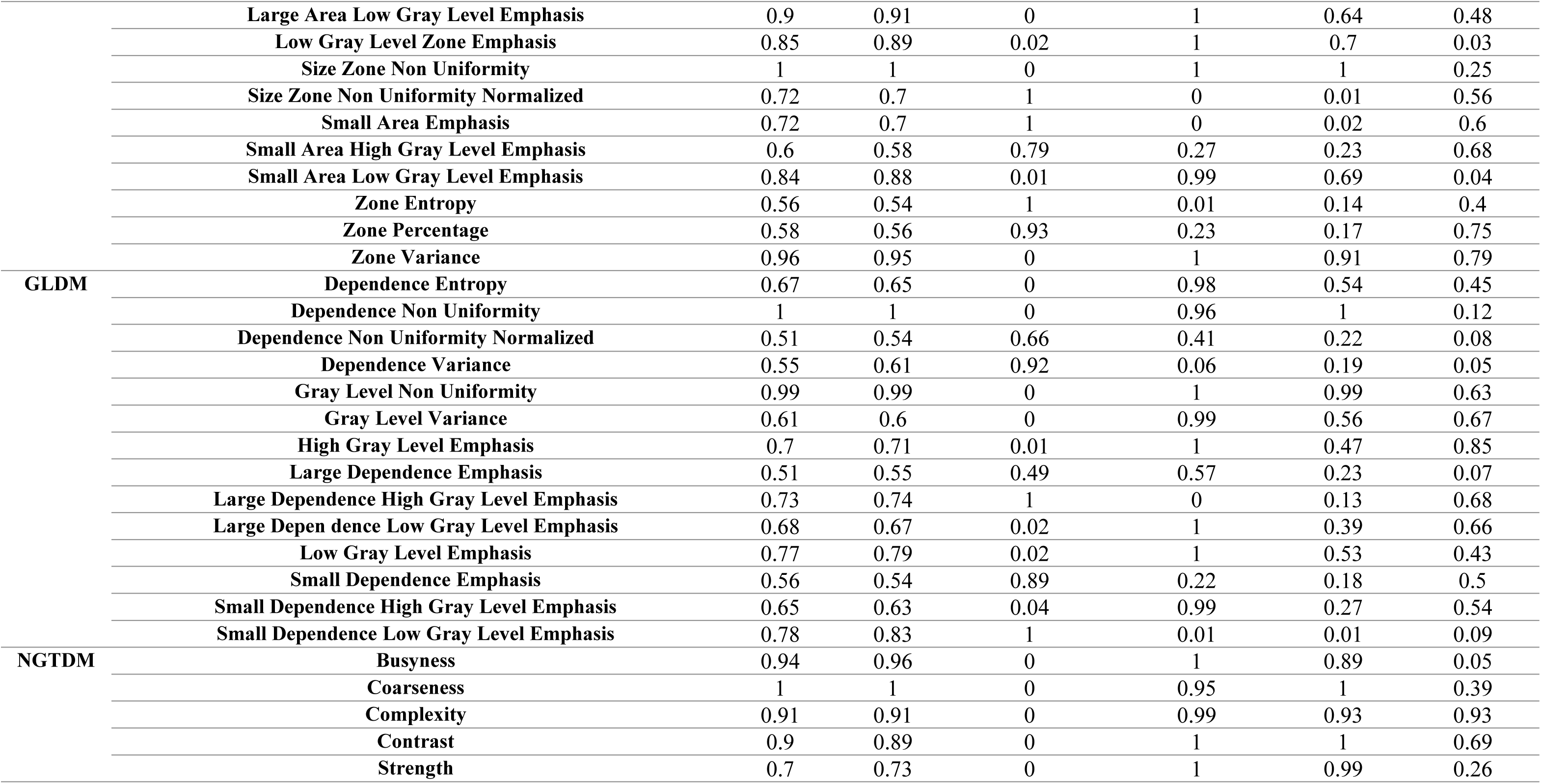
Univariate analysis in dataset 3

## REFERENCES

1. Wang, C., Horby, P.W., Hayden, F.G. & Gao, G.F. A novel coronavirus outbreak of global health concern. Lancet 395, 470 (2020).

2. La Marca, A., et al. Testing for SARS-CoV-2 (COVID-19): a systematic review and clinical guide to molecular and serological in-vitro diagnostic assays. Reprod Biomed Online 41, 483–499 (2020).

3. Chan, J.F., et al. Improved Molecular Diagnosis of COVID-19 by the Novel, Highly Sensitive and Specific COVID-19-RdRp/Hel Real-Time Reverse Transcription-PCR Assay Validated In Vitro and with Clinical Specimens. Journal of clinical microbiology 58(2020).

4. Pan, Y., et al. Potential False-Negative Nucleic Acid Testing Results for Severe Acute Respiratory Syndrome Coronavirus 2 from Thermal Inactivation of Samples with Low Viral Loads. Clinical chemistry 66, 794–801 (2020).

5. Corman, V.M., et al. Detection of 2019 novel coronavirus (2019-nCoV) by real-time RT-PCR. Euro surveillance : bulletin Europeen sur les maladies transmissibles = European communicable disease bulletin 25(2020).

6. To, K.K., et al. Temporal profiles of viral load in posterior oropharyngeal saliva samples and serum antibody responses during infection by SARS-CoV-2: an observational cohort study. The Lancet. Infectious diseases 20, 565–574 (2020).

7. Ke, Q., et al. A neuro-heuristic approach for recognition of lung diseases from X-ray images. Expert systems with applications 126, 218–232 (2019).

8. Kanne, J.P., Little, B.P., Chung, J.H., Elicker, B.M. & Ketai, L.H. Essentials for Radiologists on COVID-19: An Update-Radiology Scientific Expert Panel. Radiology 296, E113–e114 (2020).

9. Varble, N., et al. CT and clinical assessment in asymptomatic and pre-symptomatic patients with early SARS-CoV-2 in outbreak settings. Eur Radiol, 1–12 (2020).

10. Long, C., et al. Diagnosis of the Coronavirus disease (COVID-19): rRT-PCR or CT? European journal of radiology 126, 108961 (2020).

11. Borakati, A., Perera, A., Johnson, J. & Sood, T. Diagnostic accuracy of X-ray versus CT in COVID-19: a propensity-matched database study. BMJ Open 10, e042946 (2020).

12. Li, Y. & Xia, L. Coronavirus Disease 2019 (COVID-19): Role of Chest CT in Diagnosis and Management. AJR. American journal of roentgenology 214, 1280–1286 (2020).

13. Awulachew, E., Diriba, K., Anja, A., Getu, E. & Belayneh, F. Computed Tomography (CT) Imaging Features of Patients with COVID-19: Systematic Review and Meta-Analysis. Radiol Res Pract 2020, 1023506–1023506 (2020).

14. Yurdaisik, I. Effectiveness of Computed Tomography in the Diagnosis of Novel Coronavirus-2019. Cureus 12, e8134–e8134 (2020).

15. Kovács, A., et al. The sensitivity and specificity of chest CT in the diagnosis of COVID-19. Eur Radiol, 1–6 (2020).

16. Lambin, P., et al. Radiomics: extracting more information from medical images using advanced feature analysis. European journal of cancer (Oxford, England : 1990) 48, 441–446 (2012).

17. Yip, S.S. & Aerts, H.J. Applications and limitations of radiomics. Phys Med Biol 61, R150–166 (2016).

18. Avanzo, M., Stancanello, J. & El Naqa, I. Beyond imaging: The promise of radiomics. Phys Med 38, 122–139 (2017).

19. Hassani, C., Varghese, B.A., Nieva, J. & Duddalwar, V. Radiomics in Pulmonary Lesion Imaging. AJR Am J Roentgenol 212, 497–504 (2019).

20. Abdollahi, H., Shiri, I. & Heydari, M. Medical Imaging Technologists in Radiomics Era: An Alice in Wonderland Problem. Iran J Public Health 48, 184–186 (2019).

21. Amini, M., et al. Multi-level multi-modality (PET and CT) fusion radiomics: prognostic modeling for non-small cell lung carcinoma. Phys Med Biol 66(2021).

22. Bouchareb, Y., et al. Artificial intelligence-driven assessment of radiological images for COVID-19. Comput Biol Med 136, 104665 (2021).

23. Edalat-Javid, M., et al. Cardiac SPECT radiomic features repeatability and reproducibility: A multi-scanner phantom study. J Nucl Cardiol (2020).

24. Khodabakhshi, Z., et al. Overall Survival Prediction in Renal Cell Carcinoma Patients Using Computed Tomography Radiomic and Clinical Information. J Digit Imaging 34, 1086–1098 (2021).

25. Khodabakhshi, Z., et al. Non-small cell lung carcinoma histopathological subtype phenotyping using high-dimensional multinomial multiclass CT radiomics signature. Comput Biol Med 136, 104752 (2021).

26. Nazari, M., Shiri, I. & Zaidi, H. Radiomics-based machine learning model to predict risk of death within 5-years in clear cell renal cell carcinoma patients. Comput Biol Med 129, 104135 (2021).

27. Shayesteh, S., et al. Treatment response prediction using MRI-based pre-, post-, and delta-radiomic features and machine learning algorithms in colorectal cancer. Med Phys 48, 3691–3701 (2021).

28. Shiri, I., Abdollahi, H., Shaysteh, S. & Mahdavi, S.R. Test-retest reproducibility and robustness analysis of recurrent glioblastoma MRI radiomics texture features. Iranian Journal of Radiology (2017).

29. Shiri, I., et al. Machine learning-based prognostic modeling using clinical data and quantitative radiomic features from chest CT images in COVID-19 patients. Comput Biol Med 132, 104304 (2021).

30. Amini, M., et al. Overall Survival Prognostic Modelling of Non-small Cell Lung Cancer Patients Using Positron Emission Tomography/Computed Tomography Harmonised Radiomics Features: The Quest for the Optimal Machine Learning Algorithm. Clinical Oncology.

31. Shiri, I., et al. COVID-19 Prognostic Modeling Using CT Radiomic Features and Machine Learning Algorithms: Analysis of a Multi-Institutional Dataset of 14,339 Patients. medRxiv (2021).

32. Tang, Z., et al. Severity assessment of COVID-19 using CT image features and laboratory indices. Physics in medicine and biology (2020).

33. Wu, Q., et al. Radiomics Analysis of Computed Tomography helps predict poor prognostic outcome in COVID-19. Theranostics 10, 7231–7244 (2020).

34. Harmon, S.A., et al. Artificial intelligence for the detection of COVID-19 pneumonia on chest CT using multinational datasets. Nat Commun 11, 4080 (2020).

35. Bai, H.X., et al. Artificial Intelligence Augmentation of Radiologist Performance in Distinguishing COVID-19 from Pneumonia of Other Origin at Chest CT. Radiology 296, E156–E165 (2020).

36. Zhang, K., et al. Clinically Applicable AI System for Accurate Diagnosis, Quantitative Measurements, and Prognosis of COVID-19 Pneumonia Using Computed Tomography. Cell 181, 1423–1433.e1411 (2020).

37. Di, D., et al. Hypergraph learning for identification of COVID-19 with CT imaging. Medical image analysis 68, 101910 (2020).

38. Xie, C., et al. Discrimination of pulmonary ground-glass opacity changes in COVID-19 and non-COVID-19 patients using CT radiomics analysis. European journal of radiology open 7, 100271 (2020).

39. Albahli, S. & Yar, G. Fast and Accurate COVID-19 Detection Along With 14 Other Chest Pathology Using: Multi-Level Classification. Journal of medical Internet research (2021).

40. Das, D., Santosh, K.C. & Pal, U. Truncated inception net: COVID-19 outbreak screening using chest X-rays. Physical and engineering sciences in medicine 43, 915–925 (2020).

41. Wang, J., et al. Prior-attention residual learning for more discriminative COVID-19 screening in CT images. IEEE transactions on medical imaging 39, 2572–2583 (2020).

42. Roberts, M., et al. Common pitfalls and recommendations for using machine learning to detect and prognosticate for COVID-19 using chest radiographs and CT scans. Nature Machine Intelligence 3, 199–217 (2021).

43. Tizhoosh, H.R. & Fratesi, J. COVID-19, AI enthusiasts, and toy datasets: radiology without radiologists. European radiology 31, 3553–3554 (2021).

44. Mongan, J., Moy, L. & Charles E. Kahn, J. Checklist for Artificial Intelligence in Medical Imaging (CLAIM): A Guide for Authors and Reviewers. Radiology: Artificial Intelligence 2, e200029 (2020).

45. Clark, K., et al. The Cancer Imaging Archive (TCIA): maintaining and operating a public information repository. Journal of digital imaging 26, 1045–1057 (2013).

46. Morozov, S., et al. Mosmeddata: Chest ct scans with covid-19 related findings dataset. arXiv preprint arXiv:2005.06465 (2020).

47. Colak, E., et al. The RSNA Pulmonary Embolism CT Dataset. Radiol Artif Intell 3, e200254 (2021).

48. Prokop, M., et al. CO-RADS: A Categorical CT Assessment Scheme for Patients Suspected of Having COVID-19-Definition and Evaluation. Radiology 296, E97–e104 (2020).

49. Shiri, I., et al. COLI-Net: Deep learning-assisted fully automated COVID-19 lung and infection pneumonia lesion detection and segmentation from chest computed tomography images. Int J Imaging Syst Technol, in press (2021).

50. van Griethuysen, J.J.M., et al. Computational Radiomics System to Decode the Radiographic Phenotype. Cancer research 77, e104–e107 (2017).

51. Zwanenburg, A., et al. The image biomarker standardization initiative: standardized quantitative radiomics for high-throughput image-based phenotyping. Radiology 295, 328–338 (2020).

52. Robin, X., et al. pROC: an open-source package for R and S+ to analyze and compare ROC curves. BMC bioinformatics 12, 1–8 (2011).

53. Pedregosa, F., et al. Scikit-learn: Machine learning in Python. the Journal of machine Learning research 12, 2825–2830 (2011).

54. Fang, X., Li, X., Bian, Y., Ji, X. & Lu, J. Radiomics nomogram for the prediction of 2019 novel coronavirus pneumonia caused by SARS-CoV-2. Eur Radiol 30, 6888–6901 (2020).

55. Tan, H.B., et al. The study of automatic machine learning base on radiomics of non-focus area in the first chest CT of different clinical types of COVID-19 pneumonia. Scientific reports 10, 18926 (2020).

56. Yousefzadeh, M., et al. ai-corona: Radiologist-assistant deep learning framework for COVID-19 diagnosis in chest CT scans. PloS one 16, e0250952 (2021).

57. Ni, Q., et al. A deep learning approach to characterize 2019 coronavirus disease (COVID-19) pneumonia in chest CT images. Eur Radiol 30, 6517–6527 (2020).

58. Zeng, Q.Q., et al. Radiomics-based model for accurately distinguishing between severe acute respiratory syndrome associated coronavirus 2 (SARS-CoV-2) and influenza A infected pneumonia. MedComm (2020).

59. Bae, J., et al. Predicting Mechanical Ventilation Requirement and Mortality in COVID-19 using Radiomics and Deep Learning on Chest Radiographs: A Multi-Institutional Study. ArXiv (2020).

60. Chandra, T.B., Verma, K., Singh, B.K., Jain, D. & Netam, S.S. Coronavirus disease (COVID-19) detection in Chest X-Ray images using majority voting based classifier ensemble. Expert systems with applications 165, 113909 (2021).

61. Amyar, A., Modzelewski, R., Li, H. & Ruan, S. Multi-task deep learning based CT imaging analysis for COVID-19 pneumonia: Classification and segmentation. Computers in biology and medicine 126, 104037 (2020).

62. Chen, H., et al. Auxiliary Diagnosis for COVID-19 with Deep Transfer Learning. Journal of digital imaging, 1–11 (2021).

63. Chao, H., et al. Integrative analysis for COVID-19 patient outcome prediction. Medical image analysis 67, 101844 (2020).

64. Chassagnon, G., et al. AI-driven quantification, staging and outcome prediction of COVID-19 pneumonia. Medical image analysis 67, 101860 (2020).

65. Lassau, N., et al. Integrating deep learning CT-scan model, biological and clinical variables to predict severity of COVID-19 patients. Nature communications 12, 1–11 (2021).

